# Microbiota Predict Infections and Acute Graft-Versus-Host Disease after Pediatric Allogeneic Hematopoietic Stem Cell Transplantation

**DOI:** 10.1101/2023.02.09.23285664

**Authors:** Elisa B Margolis, Gabriela Maron Alfaro, Yilun Sun, Ronald H Dallas, Kim J Allison, Jose Ferrolino, Hailey S Ross, Amy E Davis, Qidong Jia, Paige Turner, Victoria Mackay, Cara E. Morin, Brandon M Triplett, Li Tang, Randall T Hayden

**Affiliations:** Department of Infectious Diseases, St Jude Children’s Research Hospital, Memphis, TN, USA; Department of Biostatistics, St Jude Children’s Research Hospital, Memphis, TN, USA; Department of Bone Marrow Transplantation and Cellular Therapy, St Jude Children’s Research Hospital, Memphis, TN, USA; Department of Computational Biology, St Jude Children’s Research Hospital, Memphis, TN, USA; Department of Pathology, St Jude Children’s Research Hospital, Memphis, TN, USA; Department of Pediatrics, University of Tennessee Health Sciences Center, Memphis, TN, USA; Department of Microbiology Immunology & Biochemistry, University of Tennessee Health Sciences Center, Memphis, TN, USA; Division of Radiology and Medical Imaging, Cincinnati Children’s Hospital, Cincinnati, OH, USA

**Author notes:** Corresponding author (address, email): 262 Danny Thomas Pl MS 320 Memphis TN 38105. Co-first author (Elisa B Margolis, Gabriela Maron Alfaro).

## Abstract

Despite intensive prophylactic and pre-emptive measures, infections remain an important cause of morbidity and mortality in pediatric recipients of allogeneic hematopoietic cell transplantation (allo-HCT). Disruption of the gut microbiota has been linked to clinical outcomes after adult allo-HCT. The objective was to evaluate whether these or differing microbiota disruptions or signatures were associated with risk of infection in pediatric allo-HCT.

In a prospective observational study, fecal samples from 74 children were collected prior to conditioning and at the time of neutrophil recovery and profiled by means of 16S ribosomal rRNA sequencing. The associations between microbiome signatures and infections or acute graft-versus-host disease (aGVHD) were examined using Cox proportional-hazards analysis.

Previously associated indices of microbiome disruption in adults, including diversity and butyrate producer frequency, did not predict infection risk in pediatric allo-HCT. Unique microbiota signatures were associated with different infections or aGVHD. A ratio of strict and facultative anaerobes (e.g. *Lachnoclostridium, Parabacteroides, Clostridium spp*.) prior to conditioning predicted likelihood of bacteremia (cox hazards ratio 3.89) in first year post HCT. A distinct ratio of oral (e.g. *Rothia, Veillonella* spp.) to colonic anaerobes (e.g. *Anaerobutyricum, Dorea, Romboutsia* spp.) at neutrophil recovery predicted likelihood of bacterial infections (cox hazards ratio 1.81) and viral enterocolitis (cox hazards ratio 1.96) through first year post transplant.

Interactions between medical interventions, pediatric hosts and microbial communities may be responsible for these consistent microbiota signatures that predict infections. A future multi-center investigation will be needed to demonstrate whether these ratios can be generalized to other pediatric cohorts.

**Key Points:** (1 or 2, <140 characters): Gut microbial communities can predict which children after HCT will have infections or acute graft versus host disease. These ratios of gut community members can be used to focus prevention and treatment on children at highest risks for complications.

## Introduction

Infections are a major cause of illness and death in pediatric patients who undergo allogeneic hematopoietic cell transplantation (allo-HCT) - a curative therapy for many pediatric patients with hematologic malignant and non-malignant disorders. Allo-HCT involves a cytotoxic conditioning regimen followed by infusion of donor hematopoietic progenitor cells. This therapy results in extensive gut mucosal damage and a period (from conditioning to engraftment) without cellular immune protection^1^. In addition, many pediatric patients experience a long period of relative immune deficiency after engraftment due to their immature cellular immune system, or to immunosuppressive therapy used for prophylaxis or treatment of graft-versus-host disease (aGVHD). The result is an increased risk for infectious complications which (when combined with toxicity) limits the success and broader applicability of allo-HCT in pediatrics^2–5^.

Among adults undergoing allo-HCT, blood stream infections, pulmonary complications, relapse, GVHD and mortality have been associated with changes in the intestinal microbiota^6–13^. This link may be from the microbiome’s role in host metabolism, immune development and intestinal barrier integrity^14–16^. In addition, the microbiome may serve as a surrogate marker for confounding risk factors that may directly influence infection risk and shape or disrupt the microbial communities within a patient. Much of the adult allo-HCT microbiome studies have focused on signals of microbiota disruption^17,18^, exposure to antibiotic agents^19^ or domination of communities by single bacterial taxon^7,20^) and their association with adverse outcomes after allo-HCT (e.g., relapse, GVHD, infection, organ toxicity).

It is unclear whether findings suggestive of a relationship between microbiota composition and outcomes of allo-HCT in adults are generalizable to pediatric allo-HCT patients. Investigations into the pediatric microbiome and allo-HCT have been descriptive or limited in size^21–28^. One reason why generalizability should be questioned is the difference in composition of pediatric microbiomes compared to their adult counterparts^29–32^. Complication rates may also differ in pediatric allo-HCT patients, with infections remaining the most common complication. Immune reconstitution, which influences complications (e.g. infections, GVHD, recurrence), may also differ between adult and pediatric patients^16^. Many of the factors that determine microbiota composition and resilience to disruption-nutrition, antibiotic regimens, microbiota temporal stability and conditioning regimens-differ widely between adult and pediatric transplantation centers^33,34^. Finally, there are large differences in pediatric immune systems (e.g. thymic function, T cell maturity and T cell diversity) compared to those of adults which contribute to differences in both microbiome and in frequency of allo-HCT complications in the two populations. Here we investigate microbiota composition, phylotypes and disruption to identify associations with clinical outcomes among pediatric recipients of allo-HCT.

## Methods

### Subjects

Stool samples and clinical data were prospectively collected from pediatric patients 18 years or younger undergoing allo-HCT at St. Jude Children’s Research Hospital between 2016 and 2019. Patients received anti-infective prophylaxis and empiric therapy per institutional guidelines (Supplemental Materials 1.1). The study protocol was approved by the institutional review board (IRB). Written informed consent and assent were obtained from all participants per institutional guidelines. Deidentified leftover stool samples from age matched healthy children collected at Seattle Children’s Hospital for other previous research studies were used for comparison. The use of these samples was designated as non-human research by the Seattle Children’s and St. Jude IRBs.

### Microbiome

DNA extraction, polymerase-chain-reaction amplification of 16S rRNA V3-V4 regions and sequencing were performed as described in the Supplementary Materials (1.3). Sequence variants were generated using DADA2 (ver 1.12.0)^35^, and then phylogenetically mapped to a custom reference set generated from full length 16S rRNA gene sequences from Ribosomal Database Project (Taxonomy 16)^36^, previously validated^37^. Phylotypes were formed by grouping amplicon sequence variants into clusters based on phylogenetic distance^38^.

Four microbiome community indices that may indicate disturbance were investigated (Supplemental Materials 1.5): diversity, bacterial load, relative frequency of butyrate producing bacteria and dominance. Diversity (evenness and richness of bacterial taxons within a community) was measured as the Balanced Weighted Phylogenetic Diversity (BWPD)^39^. As microbial gut diversity in children is strongly linked to age, communities were stratified into higher-diversity and lower-diversity groups according to the 20^th^ percentile for age matched healthy children (see SF2). Bacterial load was determined using quantitative 16S rRNA gene quantitative PCR (qPCR)^40^ and stratified based on the 20^th^ percentile for age matched healthy children (see SF3). Butyrogens were identified based on comprehensive survey^41^ using methodology described in previous studies^42^. The relative frequency of butyrogens is significantly higher in children undergoing HCT than adult studies^42,43^; stratification was based on median of age matched healthy children (see SF4). Dominance measures the relative frequency of the most abundant taxa and is also more prevalent in young children than adult gut microbiomes, higher-dominance and lower-dominance groups were stratified according to the 90^th^ percentile of abundance for age matched healthy microbiome (see SF5). None of these indices provide any information about bacterial taxa composition.

Compositional attributes were considered in terms of relative ratios of phylotypes using a new method that allowed for greater variation in microbiomes. This novel method yielded ratios composed of phylotypes hypothesized to have equivalent roles in variable microbiomes. These ratios, similar to ANCOM^44^, allowed for capturing compositional variation (and ecological equivalence) while normalizing for sequence depth and bacterial load. Specific ratios were constructed of screened phylotypes that differ between groups in abundance and variation identified by Corncob (release 0.2.0)^45^ with a false discovery rate (FDR) of 0.00001, as described in the Supplementary Materials (1.6). The ratios were constructed with a small constant (1) added to denominator and numerator read counts to account for excess zeros common in microbiome communities. Ratios screened and associated sequence variants of phylotypes are available in Supplementary Materials (ST1 and ST6).

### Outcomes

The primary outcome was the number of Microbiologically Defined Infections (MDI), which required positive laboratory result associated with clinical symptoms. Definitions for specific infection definitions can be found in Supplementary Materials (1.7). MDIs included bacteremia, fungemia, urinary tract infections (UTI), viremia (AdV, CMV, EBV, HHV6, BK virus), *Clostridioides difficile* infection (CDI), enterocolitis (AdV, rotavirus, norovirus or Cryptosporidium), skin or mucosa lesions positive for HSV and lower respiratory tract infection (e.g. bacterial pneumonia, pulmonary aspergillus or viral LRTI). Secondary outcomes included aGVHD (of any grade), bacteremia, CDI and viral enterocolitis.

### Statistics

Association between microbial community indices (diversity, bacterial load, butyrogens and dominance) or phylotype ratios and outcomes were assessed with overall Cox proportional-hazards multivariable regression models. Analyses of clinical outcomes are presented as hazard ratios with 95% confidence intervals. For infection outcomes, adjustments in multivariable models were made for age, gender and conditioning regimen (myeloablative or reduced intensity/non-myeloablative). For aGVHD, additional adjustments in multivariable models were made for graft manipulation (T-cell depletion) and graft source (apheresis or bone marrow). To compare continuous and categorical microbiota features between groups a Wilcoxon rank sum test and Fisher’s exact test were used respectively. Microbiota composition was visualized with the t-distributed stochastic neighbor embedding (t-SNE) algorithm (Supplementary Materials 1.8).

### Data Sharing Statement

All microbiome sequence data have been deposited in the National Center for Biotechnology Short Read Archive under accession number PRJNA891765.

## Results

### Characteristics of the Patients

Seventy-four pediatric patients undergoing allo-HCT at St Jude Children’s Research Hospital, who had stool samples collected prior to conditioning and at the time of engraftment were included in this cohort. Patient characteristics are described in Table 1. Leukemia was the most common indication for transplantation. Mobilized peripheral blood progenitor cells were the most common graft type with *ex-vivo* T cell-depletion in half of grafts. The most common infections were viral enterocolitis, CDI and bacteremia, each occurring in more than 30% of the participants. Severe aGVHD (grade 3-4) was rare and occurred in less than 20% of participants. The large range in age, conditioning regimens, antibiotic exposures, and graft sources contributed to considerable heterogeneity in the microbiomes of participants (Supplemental Materials Figure SF6).

**Table 1.**
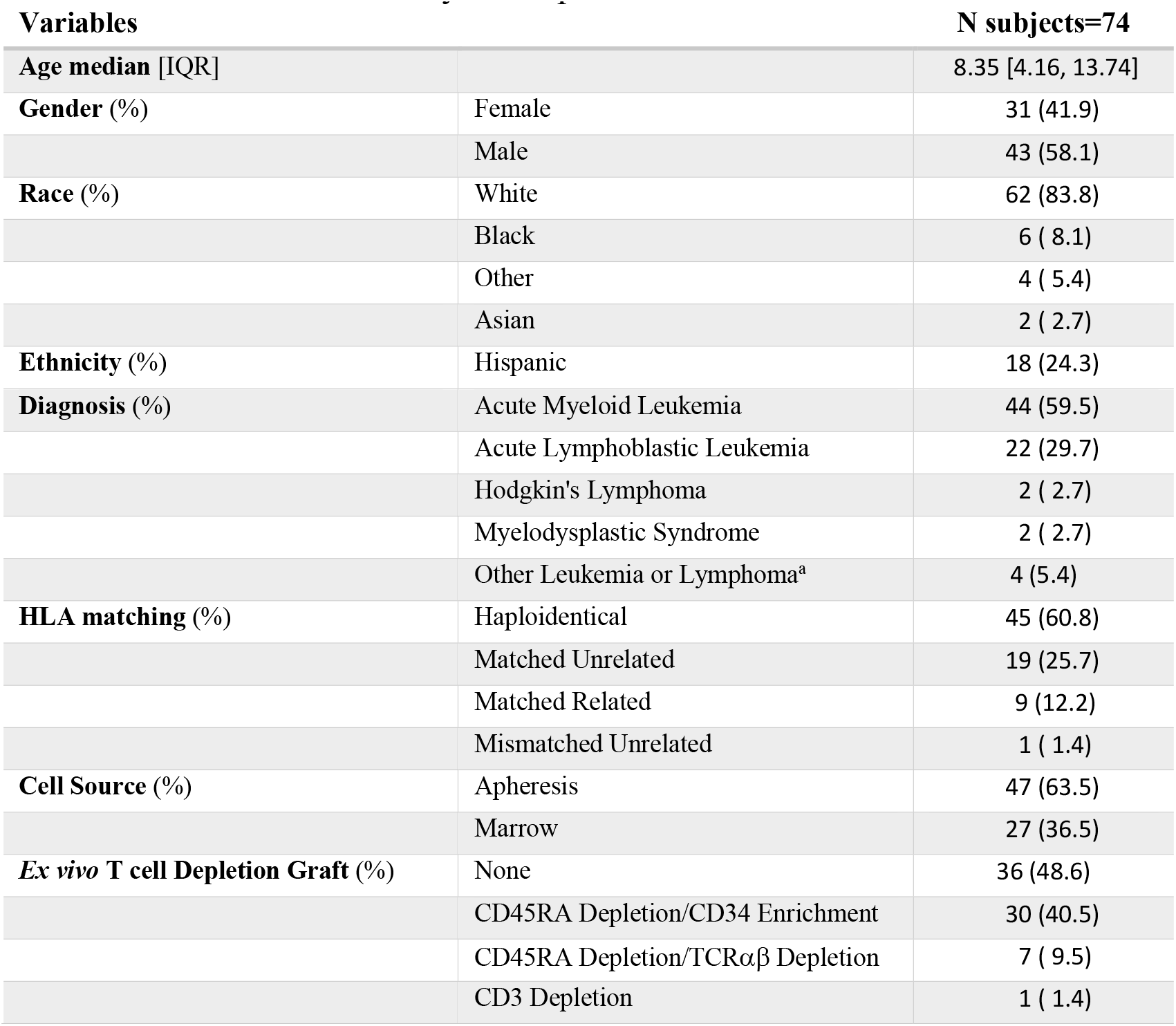
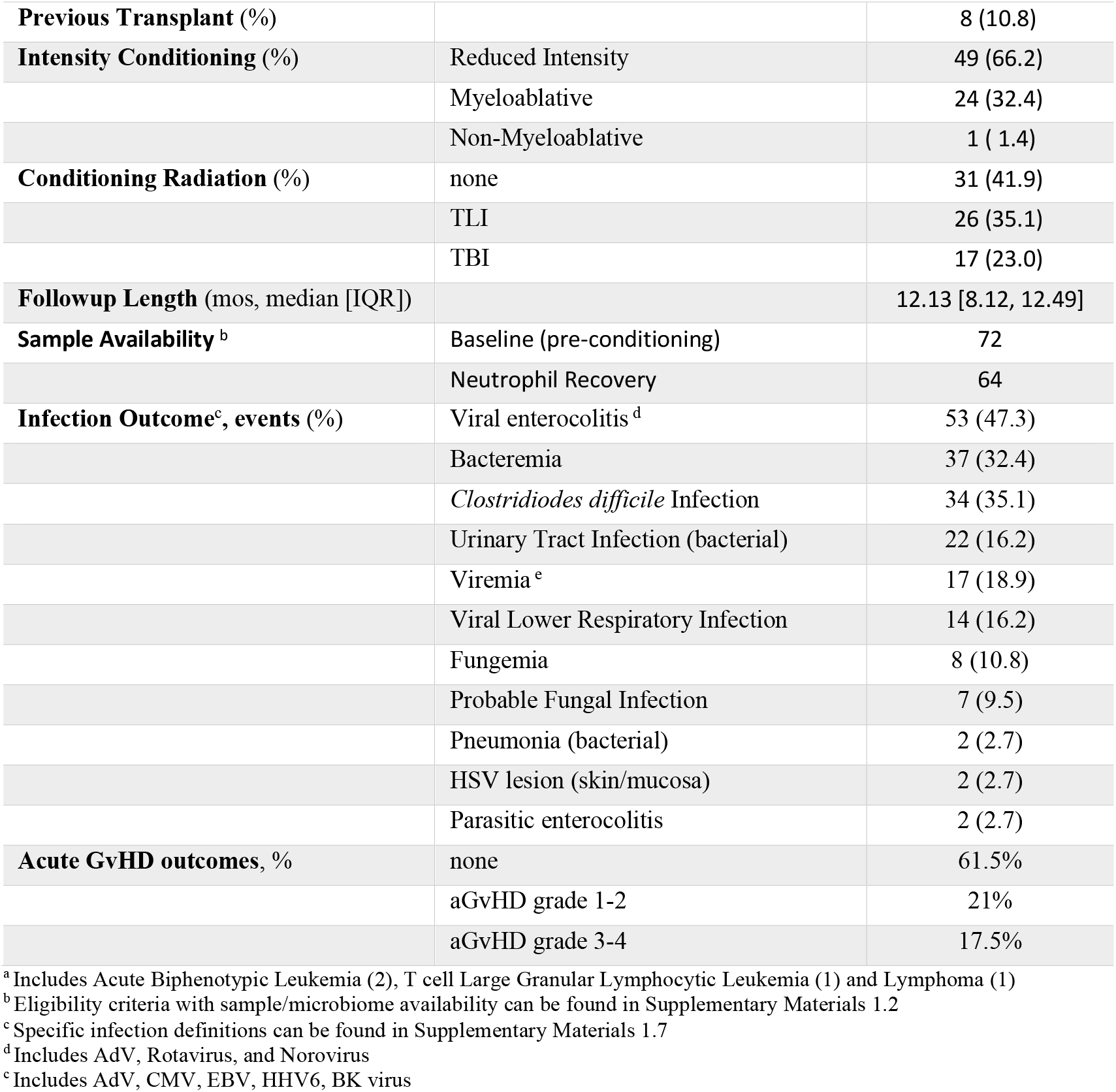
Characteristics of Study Participants

### Microbiome Disruption Indices

To characterize patterns of microbiota state in pediatric allo-HCT patients, we used 16S ribosomal RNA gene sequencing to analyze serial stool samples that had been collected prior to conditioning and subsequently, when neutrophil recovery had occurred. We investigated microbiome indices previously described as associated with adult allo-HCT outcomes and found that these indices were strongly dependent on patients’ age. To ensure appropriate benchmarking, we stratified these indices to age matched healthy children. In contrast to adults, we did not find any association between low diversity (less than 20^th^ percentile of healthy children), low butyrogens (less than median of healthy children) or low bacterial load (less than 20^th^ percentile of healthy children) and infectious outcomes (see Supplementary Materials ST3).

### Spectrum of Microbiome Compositional Attributes in Pediatric Transplantation

As all the microbiomes from individuals varied widely both at baseline and neutrophil recovery, we visualized these differences using the t-SNE algorithm (with each point representing a single stool sample and similar samples located closer to each other than more distinct microbiomes), see Figure 1. There was no cluster of microbiota that discriminated the number of infections; diversity was not associated with a particular composition. Other markers of microbiome disturbance (bacterial load, butyrogens, dominance) did not discriminate between samples (Supplementary Materials SF7).

**Figure 1:**
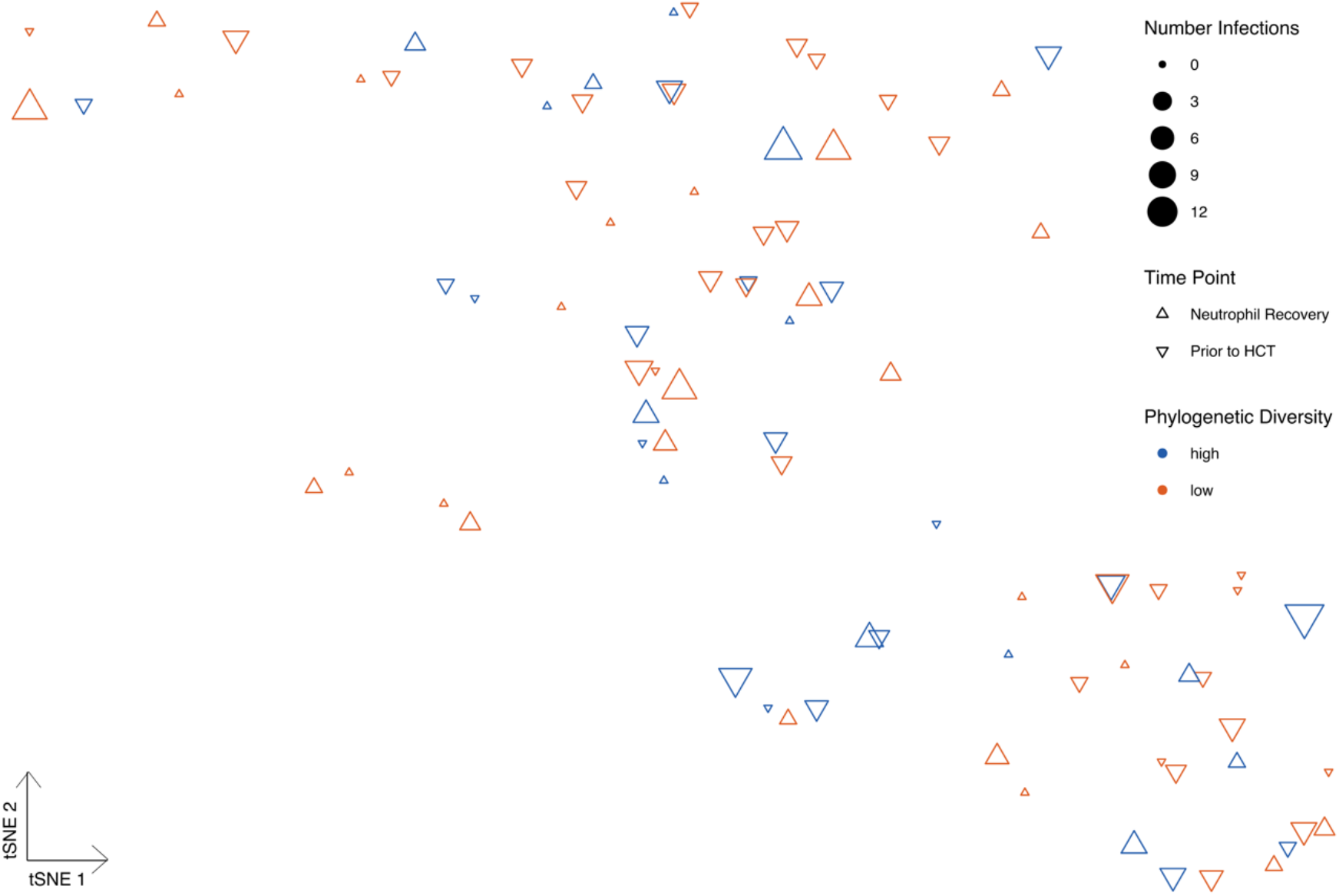
Microbiome disturbance does not predict infections in pediatric HCT. Microbiomes from 74 pediatric patients prior to conditioning and at neutrophil recovery are displayed according to their similarity (Kantorovich-Rubinstein distance) and colored based on whether phylogenetic diversity (BWPD) is lower (red) or higher (blue) than 20^th^ percentile for healthy age matched controls. The number of microbiologically defined infections determines size. Each triangle represents a single stool sample shown according to the t-distributed stochastic neighbor embedding (t-SNE) algorithm. The axes have arbitrary units with more similar samples clustering together. No particular cluster associated with either low diversity or with higher number of infections after HCT. A quantitative comparison of phylogenetic diversity, along with bacterial load, butyrate producers, domination by most abundant taxa and participants age, between samples and infections is shown in Supplemental Figure 6.

### Baseline Microbiota Predicts Specific Infections after Transplantation

Microbiota composition varied widely between individuals (Supplemental Materials SF6). To capture the unique taxa that predict outcomes, we applied a stringent screen (FDR 0.00001) for phylotypes that varied in distribution and abundance across multiple individuals^45^. These phylotypes selected to form ratios could come from similar ecological niches within their communities. These ratios were queried to determine the relationship between the baseline gut microbiota prior to transplantation and subsequent development of infection (first year after transplant). We tested each of these phylotype ratios against all outcomes (Supplemental Materials ST4). After accounting for age, gender and conditioning regimen, we found that a ratio of facultative and strict anaerobes (e.g. *Rothia sp*., *Leuconostoc sp*., *Anaaerostipes butyraticus*), predicted bacterial infections (cox hazards ratio 1.72; 95% confidence interval [1.25;2.37]). A distinct ratio of facultative and strict anaerobes (e.g. *Lachnoclostridium, Parabacteroides, Clostridium spp*.) at baseline predicted bacteremias (cox hazards ratio 3.89; 95% confidence interval [2.10;7.21]). While viral enterocolitis could be predicted (cox hazards ratio 2.76; 95% confidence interval [1.75;4.36]) based on ratio of specific butyrate producing bacteria (e.g. *Peptoniphilus, Anaerobutyricum, Blautia* spp.). Results are shown in Figure 2.

**Figure 2:**
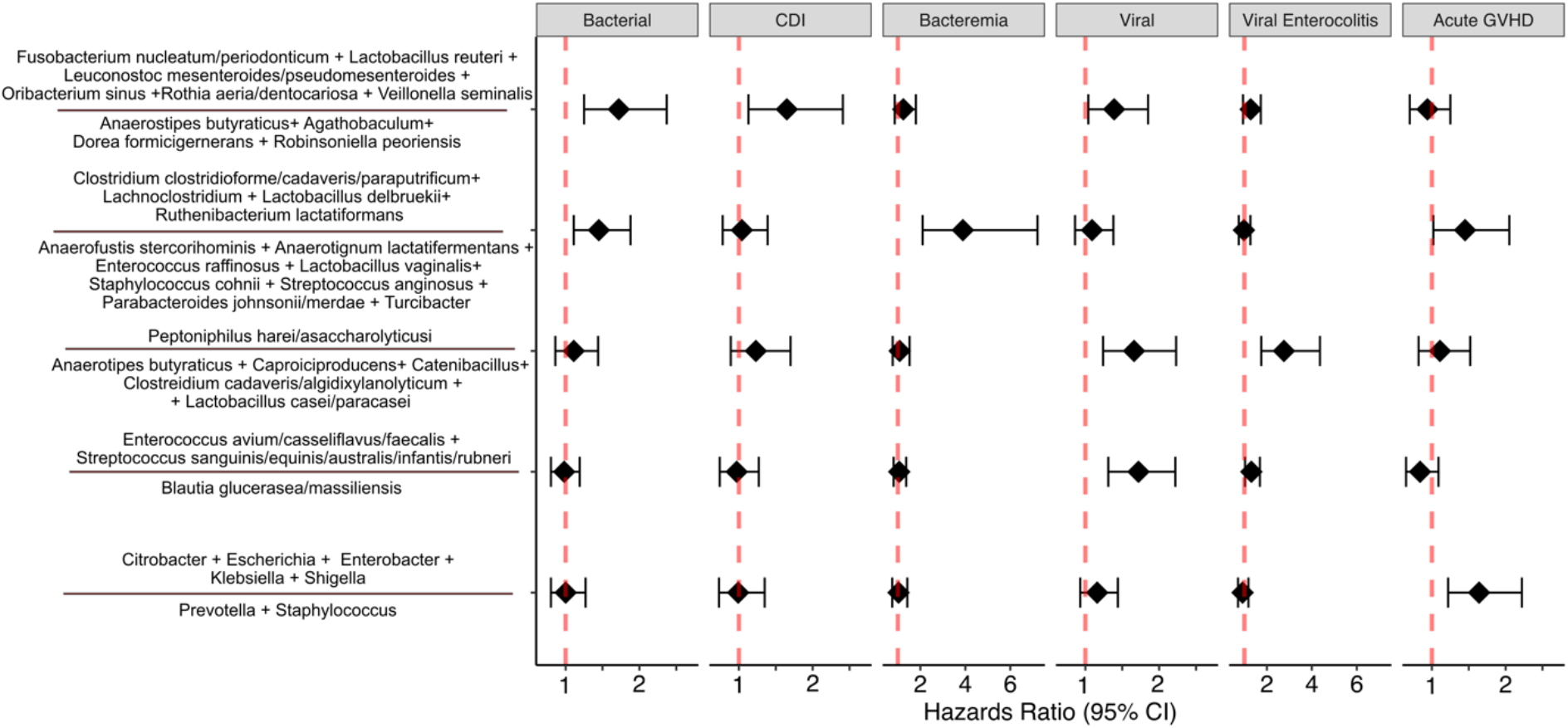
Particular ratios of bacteria species found in the microbiota of pediatric patients prior to conditioning predict infections that occur in the year after transplant. The phylotypes chosen for ratios showed similar patterns of differing in abundance and distribution in different communities. Outcomes considered included: bacterial infections (including bacteremia, pneumonia, UTI and CDI), bacteremia (alone), CDI, viral infections (included reactivation of systemic viruses, viral enterocolitis and viral LRTIs), viral enterocolitis, and aGVHD (any grade). Only microbiota ratios (log10 transformed) where the Cox Hazard Ratios for which the 95% confidence interval did not include 1, after adjusting for age, gender and conditioning regimen were considered predictive for infectious outcomes. Cox regression analysis for all 10 ratios considered is available in Supplemental Materials ST4 and quantitative differences in these ratios is available in Supplemental Materials SF8.

### Neutrophil Recovery Microbiota Predicts Specific Infections after Transplantation

The relationship between gut microbiota at neutrophil recovery and subsequent development of infection (from engraftment through first year of transplant) was characterized with the same phylotype ratios -as described above. Surprisingly none of the phylotype ratios, that were predictive of infections in baseline microbiota, were predictive at time of neutrophil recovery (Supplemental Materials ST5). However, after accounting for age, gender and conditioning regimen, we found that at neutrophil recovery a distinct ratio of oral anaerobes (e.g. *Veillonella, Rothia spp*.) and gut anaerobes (e.g. *Romboutsia timonesi, Anaerobutyricum hallii*) was predictive of bacterial infections (cox hazards ratio 1.81; 95% confidence interval [1.30;2.52]) and viral enterocolitis infections (cox hazards ratio 1.96; 95% confidence interval [1.38;2.79]). Another ratio of aerotolerant bacteria (e.g. *Klebsiella pneumoniae, Ruminococcus gnavus*) and strict anaerobes (e.g. *Bacteroides xylanisolvens, Lactobacillus gasseri*) was predictive of bacteremia (cox hazards ratio 2.25; 95% confidence interval [1.33;3.78]) at neutrophil recovery (Figure 3).

**Figure 3:**
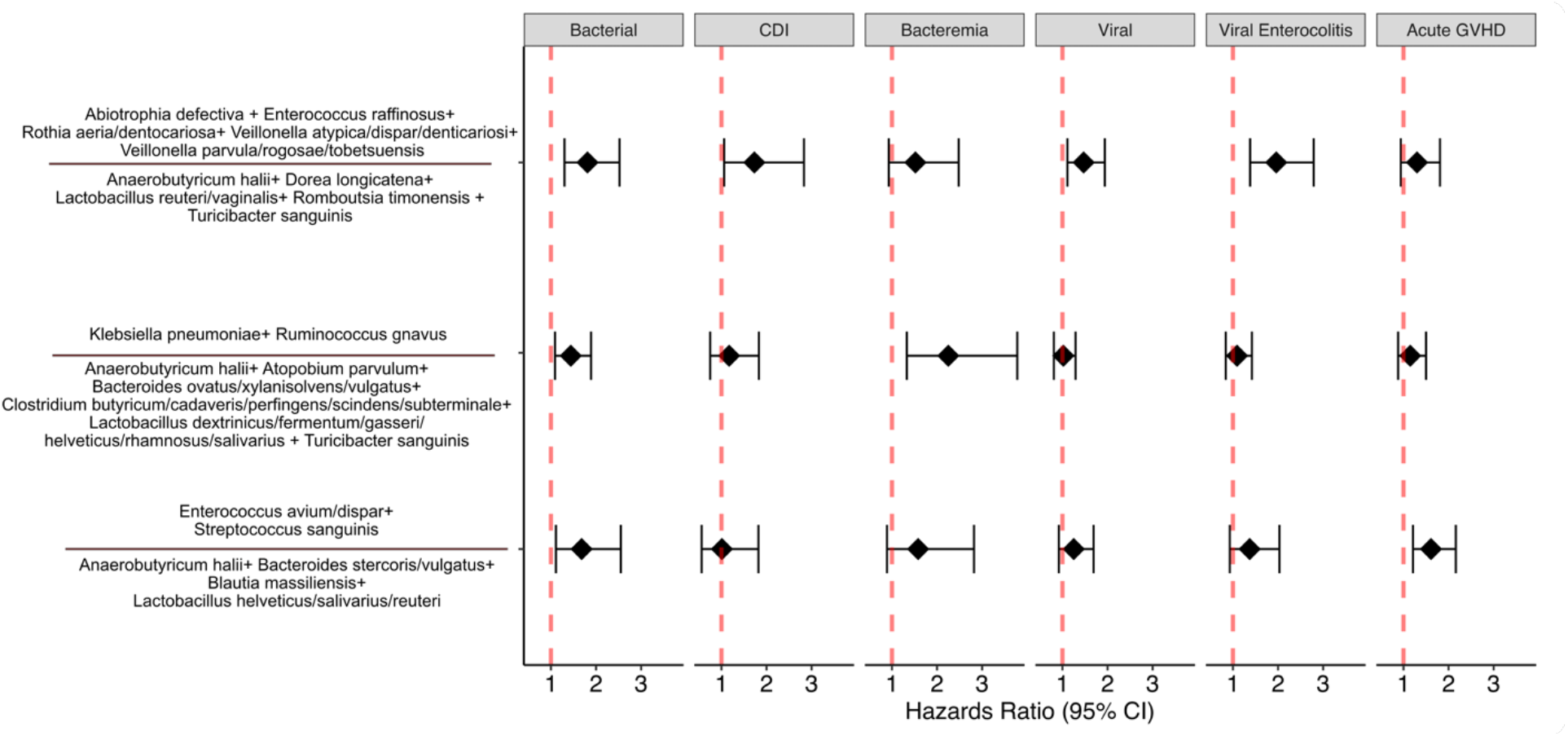
Distinct ratios of bacteria species found in the microbiota of pediatric patients at time of neutrophil recovery predict infections that occur through the first year of transplant. The phylotypes chosen for ratios showed similar patterns of differing in abundance and distribution in different communities. Outcomes considered included: bacterial infections (including bacteremia, pneumonia, UTI and CDI), bacteremia (alone), CDI, viral infections (included reactivation of systemic viruses, viral enterocolitis and viral LRTIs), viral enterocolitis, and aGVHD (any grade). Only microbiota ratios (log10 transformed) where the Cox Hazard Ratios for which the 95% confidence interval did not include 1, after adjusting for age, gender and conditioning regimen were considered predictive for infectious outcomes. Cox regression analysis for all 10 ratios considered is available in Supplemental Materials ST5 and quantitative differences in these ratios is available in Supplemental Materials SF9.

### Microbiota Signatures at Baseline and Neutrophil Recovery Predict Acute GVHD

The relationship between the gut microbiota at baseline or neutrophil recovery and subsequent development of aGVHD was characterized. We found that ratios of Enterobacter species to oral microbes (e.g. *Prevotella* and *Staphylococcus sp*.) were predictive of aGVHD (any grade) at baseline (cox hazards ratio 1.65; 95% confidence interval [1.22;2.23], see Figure 2). While at neutrophil recovery a ratio of lactate producers (e.g. *Enterococcus dispar, Streptococcus sanguinis*) to strict anaerobes (e.g. *Blautia massiliensis, Bacteroides vulgatus)* was predictive of aGVHD (any grade, cox hazards ratio 1.61; 95% confidence interval [1.21;2.16], Figure 3).

## Discussion

We found that the intestinal microbiota in pediatric patients undergoing allo-HCT are highly variable but that compositional signatures predict both infectious and aGVHD outcomes. Despite prophylaxis and pre-emptive treatment both infection and aGVHD remain serious side effects of allo-HCT in pediatric recipients. In this study, particular phylogenetic ratios reflected the microbiota composition prior to conditioning and at neutrophil recovery that were associated with these clinical outcomes.

### Unique-ness of pediatric microbiomes

It is striking that purported indicators of microbiota disruption (loss of diversity, domination by single taxa, low bacterial load and loss of butyrogens) were neither as prominent in this pediatric cohort nor linked to outcomes, as has been the case in adult allo-HCT cohorts)^6,7^. This may be due to age specific differences in what shapes microbiota (e.g. stronger founding effects, abundant butyrogens) in children or differences in resilience (e.g. adapted to frequent changes in diet, developing immune systems and more frequent antibiotic courses). Differences in methodology, such as using more sophisticated indices of diversity (e.g. phylogenetic diversity) or stratification by age, may also account for the lack of association. Another possibility is that these indices in adults are identifying a subset of allo-HCT patients at high risk for poor outcomes for which a corresponding population doesn’t exist in pediatric allo-HCT due to differences in clinical variables (e.g. indications for transplantation; prior exposures to antimicrobials, chemotherapy, or radiation; intensity of conditioning regimen; T cell depletion of grafts). Many larger-scale observational studies of the microbiome have detailed the interactions between microbial communities and pediatric allo-HCT ^16,21,28^, but few have examined outcomes^24,25^.

### Significance of Phylotype Ratios and Infection Outcome

We found only one phylotype ratio of oral anaerobes (e.g. *Veillonella, Rothia spp*.) and gut anaerobes (e.g. *Romboutsia timonesi, Anaerobutyricum hallii*) at neutrophil recovery which was able to predict cumulative infections. While the viral, bacterial, fungal and parasitic infections pathogeneses are distinct, there are multiple shared exposures (e.g. delayed immune reconstitution, breakdown of gut mucosal barrier) that can lead to increased exposure/susceptibility to these infections. It is unclear if this ratio denotes a microbiome state that alters host response to infection or results from the shared exposures and disturbances which increase susceptibility to infections. The common thread for all the infections that were predicted by a distinct microbiome signature (e.g. viral enterocolitis, bacteremia) was loss of gut epithelial architecture. There are already a few proposed mechanisms by which gut microbiome can help maintain the integrity of the mucosal barrier including links with short chain fatty acid production and tight junction assembly^46,47^, shaping mesenteric lymph nodes and Peyer’s patches^48^ and determining thickness of colon mucus^49^. An alternative is that these microbiota signatures represent expansion of microbes which have a competitive advantage in a damaged epithelial state. Interrogating how communities respond to changes induced by conditioning, radiation and antibiotics should be integrated with investigations into the components of these signatures that contribute to protective impact on maintaining the integrity of the mucosal barrier.

### Distinct Microbiota Signatures at different time points

Prior to conditioning and at neutrophil recovery are two time-points where interventions might be feasible. We found that a microbiota signature that predicts infections or aGVHD at one time point did not predict any outcomes at the other time point. This could be due to differences in the type and pathogenesis of early infections versus later infections over the time-course of pediatric allo-HCT. It is unclear if the predictive value of the baseline time point is more indicative of the composition at that time or the compositions response (or resilience) to conditioning and antibiotic course while neutropenic. Another possibility is that microbiota-host relationships may be different before and after gut injury. This is supported by investigations of butyrate producing bacteria being beneficial before but inhibiting healing after injury. This highlights that the nature of any intervention on the microbiome would require careful consideration of timing. Longitudinal studies will be integral in determining how the relationship between microbiome, host and medical interventions influence clinical outcomes.

### Significance of phylotype ratios and acute GVHD Outcome

GVHD is a major cause of morbidity after pediatric allo-HCT and its treatment is a major risk factor for opportunistic infections. We found two microbiome signatures (one at baseline and another at neutrophil recovery) which predicted the likelihood of aGVHD. The microbiome has a complicated relationship with aGVHD, as both gut decontamination^22,26,50–52^ and highly diverse adult microbiomes^8,53,54^ have been previously suggested to be protective. Our results suggest that in pediatric patients a more specific composition may be protective (which surprisingly includes the much-maligned Enterobacteriaceae) and highlight the need to investigate the middle ground between all and none. One important distinction between pediatric and adult microbiome-GVHD associations is the reliance on peripheral tolerance in adult populations while many children still have active thymic function (central tolerance). These and other differences in the immune system (e.g. T cell maturity and diversity) may alter the role that the microbiome plays in influencing aGVHD in pediatric populations.

### Caveats and Limitations

The present cohort included a large range in age, underlying diseases, conditioning regimens, antibiotic exposures, and graft sources which contributed to considerable overall heterogeneity. Nevertheless, we were able to observe parallel microbiota signatures and associations with clinical outcome that endured after controlling for some of these confounders. Another caveat to this study is that samples were analyzed by targeted amplicon sequencing of 16S ribosomal RNA gene, which limited analysis to the bacterial component of the gut microbiome and has inherent biases within the classification methodology. In addition, multiple samples had to be excluded owing to the difficulty of obtaining and processing stool samples. Not all infectious outcomes could be individually investigated due to insufficient events occurring within this cohort. The phylotype ratios also originated from screening this cohort, so to ensure that these results are generalizable, multi-institutional studies will be necessary. Finally, the largest limitation of an observational study such as this is that only correlations can be revealed. Causative relationships will require further research with preclinical models of allogeneic transplantation, infection or GVHD to demonstrate mechanisms by which microbial communities can modulate susceptibility. Correlations may still be valuable in providing a reliable method to identify which subsets of pediatric allo-HCT patients are at highest risk for infections in order to target prophylactic and pre-emptive treatment.

### Actionable

This study highlights two timepoints- pre-conditioning and neutrophil recovery-which offer brief windows during which strategies to alter or remediate microbiota influences could be evaluated. More immediately evaluating microbiota at these time points could be used to stratify patient infection or aGVHD risk and target treatment or pre-emptive diagnostics. One advantage of this study is that we used tractable patterns of microbiota composition, which can be extended to other clinical settings beyond pediatric allo-HCT. That consistent patterns of microbiota composition were associated with clinical outcomes highlights the potential associations between medical interventions, hosts and microbial communities. In pediatric patients undergoing allo-HCT, these associations were observed across many different ages, underlying conditions, graft sources and conditioning regimens despite individual differences in baseline microbiota composition.

## Data Availability

All data produced are available online at SRA NCBI.

## Acknowledgements

We would like to acknowledge Ben Treat for helpful feedback on drafts. Without the constant support, leadership and feedback from Elaine Tuomanen this project would have been impossible.

## Authorship Contributions

EBM, GM, RHD, BMT, LT, RTH were responsible for study conception and design; provided study materials and participants. EBM, GM, RHD, KJA, JF, AD, PT, VM collected and assembled clinical and laboratory data; EBM, GM, CEM reviewed radiology data; EBM, GM, YS, RHD, JF, QS, BMT, LT analyzed and interpreted the data; All authors had access to all data and take responsibility for the integrity of the data and analysis. All authors were involved in reviewing the draft manuscript and approved the final work.

