## Supplemental Materials for "Microbiota Predict Infections and Acute Graft-Versus-Host Disease after Pediatric Allogeneic Hematopoietic Stem Cell Transplantation"

**3 Phylotypes Included in Ratios** 17

**4 Supplemental Material References** 24

### 1 Supplemental Methods

#### 1.1 Prophylaxis and Empiric Treatment

No oral decontamination or antibacterial prophylaxis was administered peri-HCT. Trimethoprim-sulfamethoxazole (TMP-SMZ) was the preferred *Pneumocystis jiroveci* Pneumonia (PJP) prophylaxis. Micafungin (during conditioning), followed by Voriconazole (on day 2 post HCT) was used for fungal prophylaxis. Patients received acyclovir prophylaxis for first 30 days if HSV serology was positive. First line empiric treatment for fever and neutropenia was cefepime monotherapy unless there was history of multi-drug resistant organism or signs of sepsis. IVIG was given weekly for levels of IgG <400mg/dL. Viral reactivation (e.g. Cytomegalovirus (CMV), Epstein-Barr Virus (EBV), Adenovirus (AdV)) was monitored weekly for first 100 days and treated preemptively with antivirals as appropriate.

#### 1.2 Eligibility Criteria

Eligibility criteria (figure SF1) included all patients less than 19 years of age who received a hematopoietic-cell transplantation at St Jude Children's Research Hospital between 2016 and 2019. Patients were excluded from a time point if there stool samples were missing, had insufficient bacterial content for sequencing or insufficient sequence depth or did not achieve neutrophil recovery.

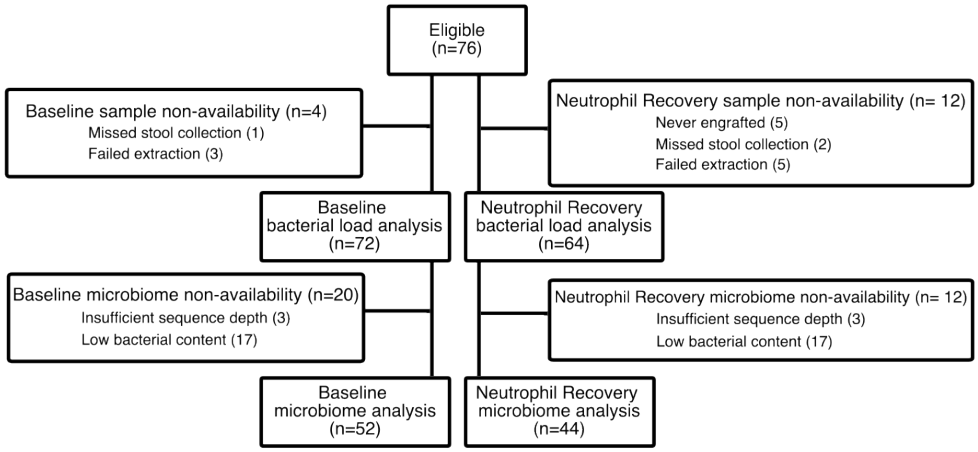

Figure SF1: Sample availability for baseline and neutrophil recovery.

#### 1.3 Sample preparation, DNA extraction, Amplicon preparation, DNA sequencing

Stool samples frozen at -80°C were diluted to 10% w/v with nuclease-free PBS and 20 mg frozen stool equivalent was extracted (modified Qiagen DNeasy PowerSoil 96 HTP (Cat. No. 1115096)) using enzyme treatment ( 10 µL of Lysozyme (10 mg/ml), 6 µL Mutanolysin (25 kU/ml), 3 µL Lyostaphin (4KU/ml) for 1 hour) and bead-beating (FastPrep) in a protocol optimized for low bacterial content stool samples[1]. Bacterial DNA content was measured using a 16S rRNA quantitative PCR using a plasmid containing E. coli 16S gene as the standard, primers and probes as previously described [2] with only change being reverse primer was changed to GGACTACHVGGGTATCTAATCCTGTT to achieve greater universality based on primers from Herlemann et al [3] . Samples with less than 1000 copies of 16S rRNA/mg stool where excluded from study. The 16S rRNA gene amplicon for sequencing used primers for the V3-V4 region[4] and a touch-down PCR protocol[5]. The PCR had 20 cycles with annealing step decreasing by 0.3C each cycle (denaturation 95°C x 20 sec, annealing range of 60-54.3°C x 15 sec and extension 72°C x 5min) followed by 20 cycles with the standard cycling conditions (denaturation 95° C x 20 sec, annealing 55°C x 15 sec and extension 72°C x 5min). Sequencing of indexed amplicons was performed on an Illlumina MiSeq instrument with miseq reagent kit v3-600 cycle to capture paired end (2x 300bp) sequences.

#### 1.4 Classification

Paired-end reads were quality filtered, derplicated and clustered into amplicon sequence variants in DADA2 (ver 1.12.0)[6]. The filtered and paired 16S rRNA gene amplicons were used to recruit full-length 16S rRNA gene sequences from Ribosomal Database Project release 16 (March 2018)[7] using vsearch (ver 2.7.1) with global alignment search. The collected reference sequences were assembled into a phylogenetic tree using RaxML (version 8.2.11)[8]; leaves with only 1 representative sequence were used to recruit additional sequences. QC-filtered amplicon sequences were placed onto the classification reference tree using the pplacer tool, using MALIAMPI[9], with the pipeline[10] validated in separate studies. To ensure that classified taxonomies were not overcalled we clustered sequence variants in terms of phylotypes based on phylogenetic distance and coabundance[9]. Note MALIAMPI allows with harmonization of classification across microbiome studies so these results can be compared to previous or future cohorts[9].

#### 1.5 Stratification of Microbiome Metrics

Various indices that describe microbiome community structure have been used to illustrate how microbiome disruptions are associated with outcomes in adult HCT studies[11, 12, 13]. All of these indices have distinct distributions in pediatric gut microbiotas than adults, therefore we have chosen to stratify based on healthy pediatric microbiota and classify based on a percentile normalized by age. The healthy pediatric microbiota cohort were from de-identified non-research samples (designated so by St Jude and Seattle Children's Institutional Review Boards) obtained as leftover control samples (siblings of patients presenting with diarrhea who had not had diarrhea within the last month) from a previous rotavirus study.

Diversity is a measure of both the evenness and richness of bacterial taxons within a community. Many adult HCT studies use Simpsons diversity index[14, 13], we opted for a phylogenetic diversity as it is less prone to error from over–classification. Given that taxonomies have inconsistent phylogenetic relationships in bacteria we have elected to use Balanced Weighted Phylogenetic Diversity (BWPD) to measure diversity in our samples. We found that phylogenetic diversity differed significantly between age categories less than 2 years of age, 2-6 years of age and greater than 5 years of age in healthy pediatric cohort (figure SF2). We opted to stratify based on the 20th percentile healthy cohorts BWPD per age category.

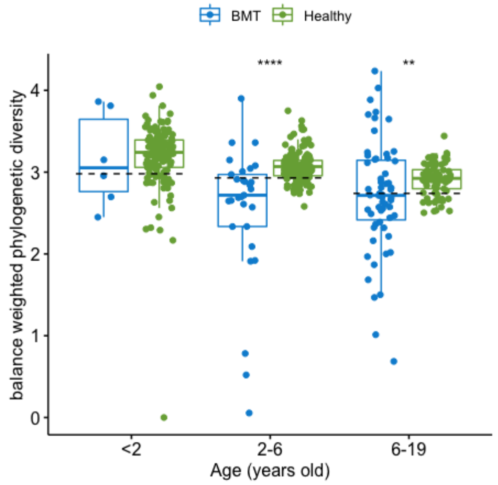

Figure SF2: Phylogenetic Diversity (BWPD) in healthy children and those in this study undergoing HCT. Twentieth percentile of healthy is demarcated with dotted lines. Significance (** $\leq$ 0.01 ; *** $\leq$ 0.001) seen between healthy cohort (green) and those in this study(blue) by Kruskal-Wallis.

Low bacterial load is the most common reason samples from HCT are excluded from microbiome studies. To determine if bacterial load, a potential indicator of disturbance, was associated with outcomes we measured the 16S rRNA gene content by quantitative PCR, as described in [2] . We found that infants (under the age of 2) had on average higher stool bacterial load in healthy cohort(figure SF3), therefore we opted for the twentieth percentile of healthy cohort bacterial load per age category.

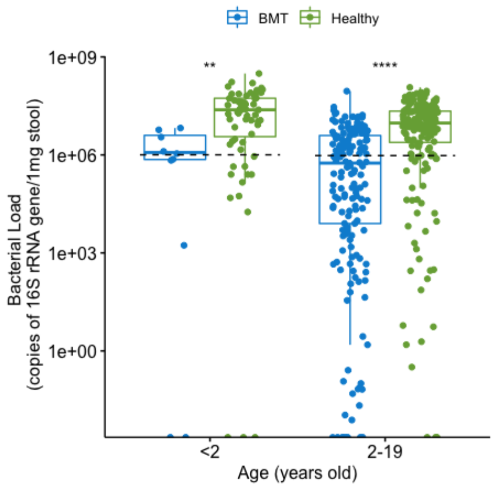

Figure SF3: Stool Bacterial Load (as measured in copies of 16S rRNA gene copies per 1mg stool) in healthy children and those in this study undergoing HCT. 20th percentile of healthy is demarcated with dotted lines. Significance (** $\leq$ 0.01 ; *** $\leq$ 0.001) seen between healthy cohort (green) and those in this study(blue) by Kruskal-Wallis.

Butyrate production is one of the key links to microbiome metabolism and host response[15, 16]. Studies have linked butyrate production to viral LRTI[11] and recovery from GVHD[17] in adult HCT. Here we infer the potential to produce butyrate based on relative frequency of known butyrate producers[18]. Known butyrate producers are more common in childhood microbiota (2-19 year olds) than that found in adults or in infants (less than 2 years old), therefore we opted to stratify based on the 50th percentile of healthy cohorts butyrogens per age category(figure SF4).

Taxon dominance is a measure of evenness of bacterial taxons within a community that looks only at the most abundant taxons relative abundance and has been associated with outcomes in adult HCT studies[12, 19]. We found that single taxon dominance is quite common in healthy infant microbiota (figure SF5), with dominance differing significantly between age categories less than 2 years of age and greater than 2 years of age. We opted to stratify based on the 90th percentile of healthy cohorts dominance per age category.

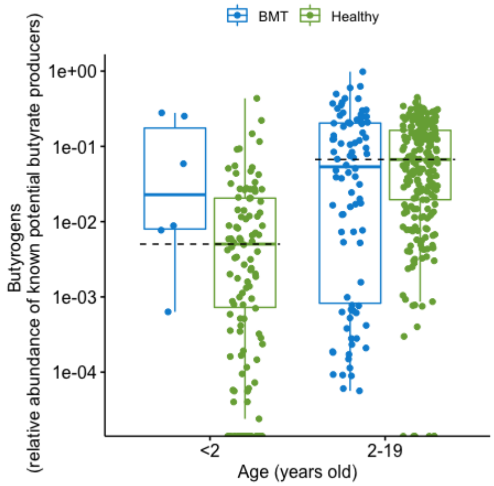

Figure SF4: Butyrogens (relative abundance of potential butyrate producing taxa) in healthy children and those in this study undergoing HCT. Fiftieth percentile healthy is demarcated with dotted lines. No significance difference seen between healthy cohort (green) and those in this study(blue) by Kruskal-Wallis.

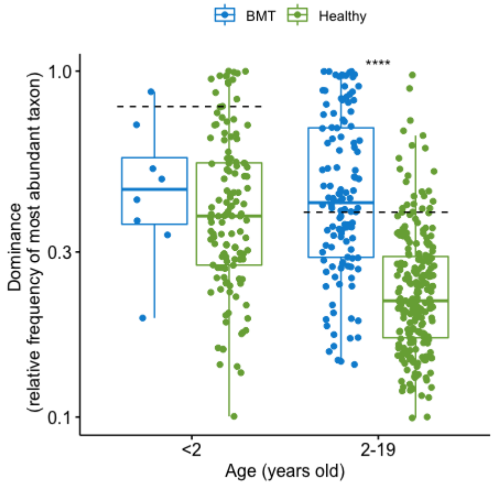

Figure SF5: Dominance (relative frequency of the most abundant taxon) in healthy children and those in this study undergoing HCT. Nintieth percentile healthy is demarcated with dotted lines. Significance (*** $\leq 0.001$) seen between healthy cohort (green) and those in this study(blue) by Kruskal-Wallis.

#### 1.6 Composition Attributes and Phylotype Ratios

As composition can vary widely between individuals within a group and important taxa may be absent in various communities, we screened for phylotypes that persistently vary in distribution and abundance between outcome groups identified by Corncob release 0.2.0 (Count Regression for Correlated Observations with the Beta binomial)[20] with a false discovery rate of 0.00001. Screened phylotypes were included in preliminary and final composition attribute ratios if variance persisted when outliers excluded (modified mean). Composition ratios of absolute abundances were constructed from screened phylotypes, with excess zeros of phylotype abundances handled with small positive constant (1) before performing read count ratio and log transformations similar in approach to ANCOM[21]. Ratios that resulted in describing similar taxonomies were discarded in favor of simpler ratios that had fewer total phylotypes. The following (table ST1) is a complete list of the ratios that were considered in this study (all ratios were tested against all outcomes). followed by the sequence variants that made up each of the phylotypes considered.

#### 1.7 Specific Infection Definitions

Bloodstream infection was defined as detection of bacteria (either recognized bacterial pathogen[22] from or $\geq 1$ blood culture or common commensal bacteria if identified from $\geq 2$ blood culture sets ) or fungi in blood cultures obtained for clinical care. Viral enterocolitis was defined as detection of adenovirus, rotavirus or norovirus by PCR in stool sample obtained with documented clinical diagnosis of diarrhea (>3 loose or watery stools in a 24-hour period). *Clostridiodes difficile* infection was defined as identification of toxin gene PCR from loose or watery stool. Parasitic enterocolitis was diagnosed by presence of Cryptosporidium antigen in loose or watery stool. Lower respiratory tract infection was defined as either evidence of infection from bronchoaveolar lavage sample (e.g. positive culture, aspergillus antigen>0.5 or positive viral PCR) or new possible or definite pulmonary infiltrate found in chest imaging (as assessed by radiologist blinded to microbiome and microbiology results of the participants) with positive upper respiratory tract sample (NP swab or tracheal aspirate)[11]. Viremia was defined as the presence of virus in blood stream that would meet treatment standards for Adenovirus (quantitative PCR above 3 log in first four months after HCT), CMV (quantitative PCR above 3 log), EBV (quantitative PCR above 4 log or rising in 3 consecutive weekly tests) or BK virus (requires quantitative PCR above 4 log in blood and above 7 log in urine)[23]. Recurrent episodes of any infection was only considered if occurred more than 14 days after initial positive bacteria culture and more than 90 days for viral infections. Only exception was no recurrence included for reactivation of systemic viruses (e.g. CMV, HHV6, EBV viremia).

#### 1.8 Statistical Methods

Primary statistical analysis between clinical outcomes and microbial ratios or indices was Cox proportional-hazards multivariable regression model performed using R package survival (coxph). When baseline samples were analyzed, outcomes were considered for all events from HCT day 0. When neutrophil engraftment samples were analyzed, outcomes from neutrophil engraftment were included (did not include day 0 to neutrophil engraftment events). All data was censored at one years of follow-up. For infection outcomes, adjustments in multivariable models were made for age, gender and conditioning regimen (myeloablative or reduced intensity/non-myeloablative). For acute GVHD, additional adjustments in multivariable models were made for graft manipulation (T-cell depletion) and graft source (apheresis or bone marrow). When community indices was considered as a binary (high v low) variable, patients were split into groups based on a percentile compared to healthy pediatric microbiota and normalized by age. We used the Rtsne package to visualize microbiota composition based on t-distributed stochastic neighbor embedding (t-SNE) dimensionality reduction. The classification down to a species level was used as input to the t-SNE algorithm.

| Name | Phylotypes | Taxon Represented |
|---|---|---|
| ratio 3.3 | pt___00189, pt___00421, pt___00235 | Abiotrophia defectiva + Enterococcus raffinosus + Rothia aeria + Rothia dentocariosa |
| | pt___00018, pt___00009, pt___00016, pt___00074, pt___00111, pt___00023, pt___00027, pt___00019, pt___00228 | Lachnospiraceae + Lactobacillus rhamnosus + Lactobacillus casei group + Lactobacillus casei+ Clostridiales + Streptococcus parasanguinis + Streptococcus australis + Ruminococcus + Ruminococcus faecis + Faecalimonas umbilicata + Lachnospiraceae + Glucerabacter + Schaalia odontolytica + Actinomyces + Schaalia+ Akkermansia muciniphila + Anaerostipes + Anaerostipes butyraticus |
| ratio 3.16 | pt___00189, pt___00421, pt___00235, pt___00001 | Abiotrophia defectiva + Enterococcus raffinosus + Rothia aeria + Rothia dentocariosa+ Veillonella parvula + Veillonella rogosae + Veillonella dispar + Veillonella + Veillonella atypica+ Veillonella tobetsuensis + Veillonella denticariosi |
| | pt___00043, pt___00003, pt___00024 , pt___00068, pt___00191, pt___00015 | Dorea longicatena + Dorea + Romboutsia timonensis + Romboutsia + Anaerobutyricum + Anaerobutyricum hallii + Lactobacillus reuteri + Lactobacillus vaginalis + Turicibacter sanguinis + Turicibacter |
| ratio 5.0 | pt___00005, pt___00021 | Klebsiella pneumoniae+ Ruminococcus gnavus |
| | pt___00025, pt___00018 , pt___00057, pt___00138, pt___00268, pt___00097, pt___00015, pt___00001, pt___00024, pt___00200, pt___00031 , pt___00054, pt___00067, pt___00198, pt___00086, pt___00065, pt___00079 , pt___00042 | Lactobacillus dextrinicus+ Lactobacillus fermentum+ Lactobacillus gasseri+ Lactobacillus helveticus+ Lactobacillus rhamnosus+ Lactobacillus salivarius+ Turcibacter sanguinis+ Veillonella dispar+ Anaerobutyricum hallii+ Atopobium parvulum+ Bacteroides xylanisolvens+ Bacteroides ovatus+ Bacteroides vulgatus+ Bacteroides ovatus xylanisolvens+ Clostridium butyricum+ Clostridium perfringens+ Clostridium scindens+ Clostridium subterminale+ Clostridium cadaveris+ Clostridium symbiosum |
| ratio 7.8 | pt___00076, pt___00033 , pt___00216, pt___00086, pt___00049, pt___00652 | Ruthenibacterium lactatiformans + Clostridium clostridioforme + Lachnoclostridium + Clostridium + Clostridium cadaveris + Clostridium paraputrificum + Lactobacillus delbrueckii |
| | pt___00421, pt___00263, pt___00706, pt___00179 , pt___00237, pt___00191, pt___00676, pt___00124, pt___00201 | Enterococcus raffinosus + Staphylococcus cohnii + Streptococcus anginosus + Parabacteroides merdae + Parabacteroides johnsonii + Turicibacter + Lactobacillus vaginalis + Clostridium + Anaerofustis + Anaerofustis stercorihominis + Anaerotignum lactatifermentans + Anaerotignum |
| ratio 9.0 | pt___00046, pt___00482 | Prevotella + Staphylococcus |
| | pt___00273, pt___00630, pt___00175, pt___00649, pt___00020, pt___00005 | Enterobacter+ Citrobacter+ Escherichia+ Escherichia/Shigella + Klebsiella |
| ratio 11.26 | pt___00235, pt___00193, pt___00301, pt___00127, pt___00068 , pt___00180 | Rothia aeria + Rothia dentocariosa + Veillonella seminalis + Veillonella + Oribacterium sinus + Fusobacterium nucleatum + Fusobacterium periodonticum + Lactobacillus reuteri + Leuconostoc mesenteroides + Leuconostoc pseudomesenteroides |
| | pt___0157, pt___00093, pt___00450, pt___00395 | Anaerostipes + Anaerostipes butyraticus + Dorea formicigenerans + Agathobaculum + Robinsoniella peoriensis |
| ratio 12.0 | pt___00163, pt___00117, pt___00022 | Enterococcus avium+ Enterococcus dispar+ Streptococcus sanguinis |
| | pt___00031, pt___00024, pt___00053, pt___00038, pt___00057, pt___00068, pt___00097 | Anaerobutyricum hallii+ Bacteroides stercoris+ Bacteroides vulgatus + Blautia massiliensis+ Lactobacillus helveticus+ Lactobacillus reuteri+ Lactobacillus salivarius |
| ratio 16.16 | pt___00234, pt___00542 | Peptoniphilus harei + Peptoniphilus asaccharolyticus |
| | pt___00086, pt___00131, pt___00264, pt___00836, pt___00047, pt___00228, pt___00285 | Clostridium + Clostridium cadaveris + Clostridium algidixylanolyticum + Catenibacillus + Caproiciproducens + Lactobacillus paracasei + Lactobacillus casei + Lactobacillus casei group + Anaerostipes + Anaerostipes butyraticus + |
| ratio 18.0 | pt___00002, pt___00039, pt___00163, pt___00007, pt___00190, pt___00009 | Enterococcus avium + Enterococcus casseliflavus + Enterococcus faecalis+ Streptococcus sanguinis+ Streptococcus equinus infantarius+ Streptococcus australis_infantis+ Streptococcus australis_rubneri |
| | pt___00038, pt___00142 | Blautia glucerasea + Blautia massiliensis |
| ratio 25.26 | pt___00054, pt___00304, pt___00288 | Bacteroides ovatus + Bacteroides xylanisolvens + Schaalia+ Schaalia turicensis+ Actinomyces |
| | pt___0157, pt___00625, pt___00093, pt___00450, pt___00395 | Anaerostipes + Anaerostipes butyraticus + Intestinimonas + Dorea formicigenerans + Agathobaculum + Robinsoniella peoriensis |

Table ST1: Phylotype ratios include compositional attributes that differed in variance and abundance between groups. All 10 ratios were investigated for their ability to predict infectious and gvhd outcomes in pediatric HCT participants from gut microbiota at baseline and neutrophil recovery.

### 2 Supplemental Data

#### 2.1 Microbiome Composition

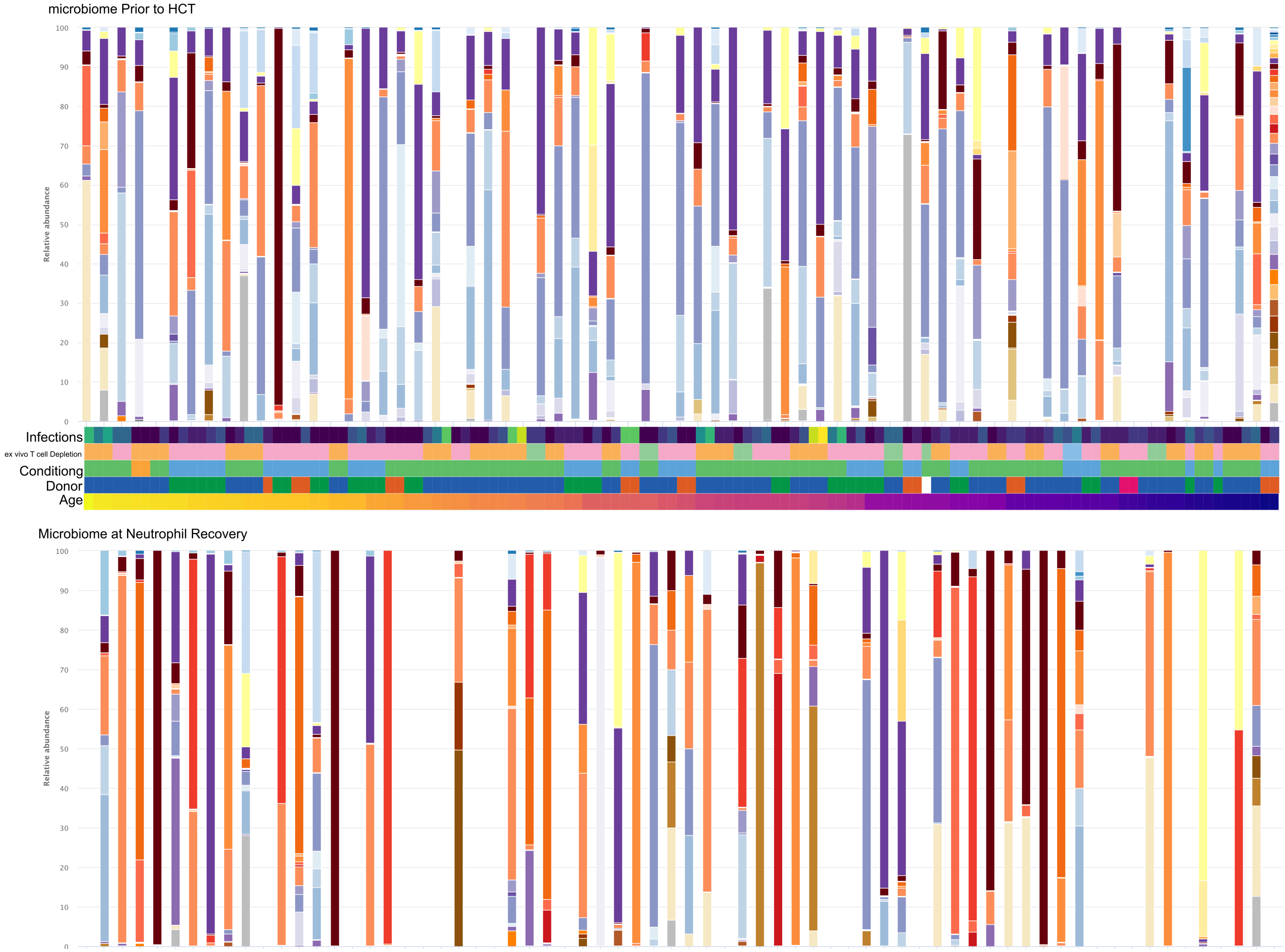

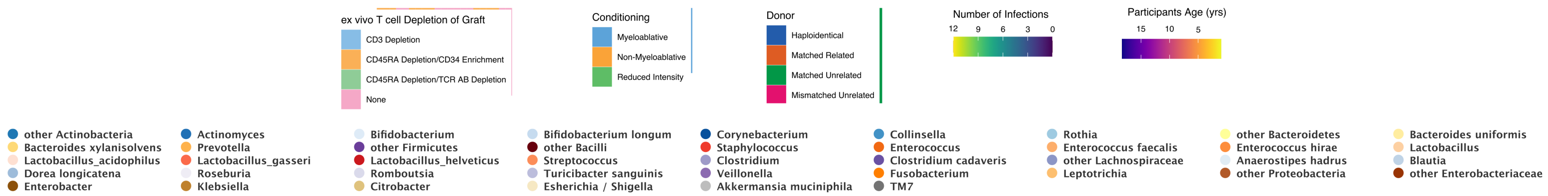

Figure SF6: Microbiome composition in 74 pediatric participants prior to HCT (above) and at neutrophil recovery (below) with each microbiome a bar displayed as relative abundance colors correspond to particular species or if not specified as other phylum/family. Participants age, donor source, conditioning regimen, T cell depletion of graft and number of infections are shown in heat map between the two microbiome charts. Participants are ordered by age on the x-axis and if available both time points are lined up.

| Indice | Category | Baseline: No MDI, Number (%) | Baseline: MDI, Number (%) | Baseline: Fischer's Exact (P Value) | Neutrophil Recovery: No MDI, Number (%) | Neutrophil Recovery: MDI, Number (%) | Neutrophil Recovery: Fischer's Exact (P Value) |
|---|---|---|---|---|---|---|---|
| Bacterial Load | | | | >0.999 | | | >0.999 |
| | high | 7 (58.3) | 34 (56.7) | | 10 (38.5) | 14 (36.8) | |
| | low | 5 (41.7) | 26 (43.3) | | 16 (61.5) | 24 (63.2) | |
| Diversity | | | | >0.999 | | | >0.999 |
| | high | 4 (40.0) | 17 (40.5) | | 6 (35.3) | 10 (40.0) | |
| | low | 6 (60.0) | 25 (59.5) | | 11 (64.7) | 15 (60.0) | |
| Single Taxa Dominance | | | | 0.255 | | | 0.754 |
| | present | 5(50.0) | 12 (26.7) | | 13 (65.0) | 21 (72.4) | |
| | absent | 5 (50.0) | 33 ( 73.3) | | 7 ( 35.0) | 8 ( 27.6) | |
| Butyrate Frequency | | | | 0.475 | | | 0.018 |
| | high | 6(60.0) | 30(71.4) | | 9(52.9) | 4(16.0) | |
| | low | 4(40.0) | 12(28.6) | | 8(47.1) | 21(84.0) | |

Table ST2: No differences observed in microbiome community indices and number of microbiologically defined infections prior to HCT (baseline) and at the time of neutrophil recovery in pediatric participants. Fischer's exact p value is adjusted for multiple comparisons.

#### 2.2 Community Indices and Number of Microbiologically Defined Infections

Gut microbiota disruption have been linked to clinical outcomes after adult HCT. The main objective or this project was to evaluate whether the same microbiota disruptions or signatures were associated with risk of infection in pediatric HCT. We investigated four signatures of disruption (i.e. Bacterial Load, Diversity, Single Taxa Dominance and Butyrate Producers Relative Frequency) and found that none were associated with having a Microbiologically Defined Infection (MDI) after HCT in pediatric samples taken prior to HCT (baseline) and at the time of neutrophil recovery (Table ST2). Further we could find no association between a particular composition cluster, these signatures of disruption, or age with the number of MDIs that occurred (Figure SF7).

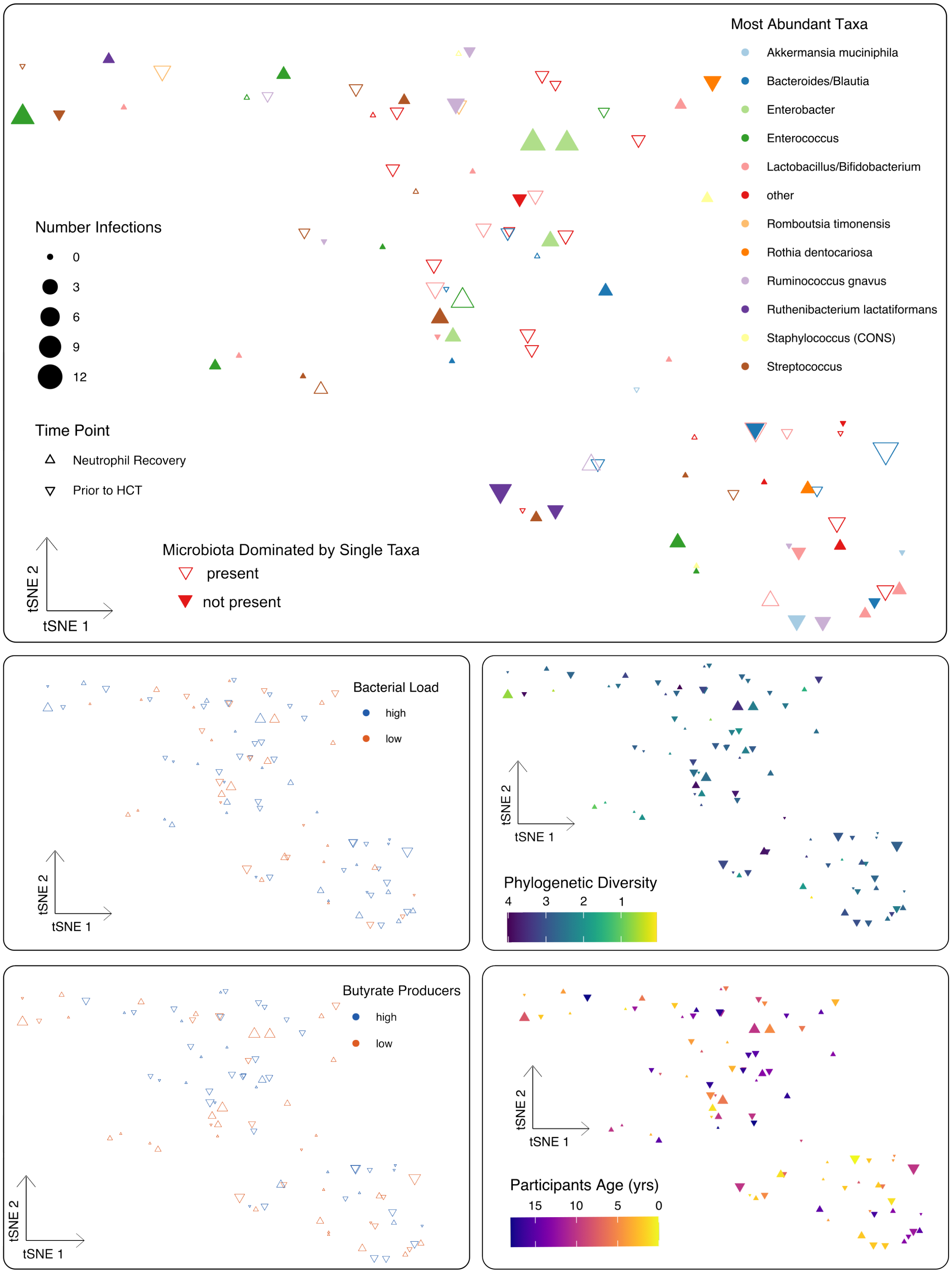

Figure SF7: Microbiome disturbance does not predict infections in pediatric HCT. Microbiomes from 74 pediatric patients prior to HCT and at Neutrophil Recovery are displayed according to their similarity (Kantorovich-Rubinstein distance). The number of microbiologically defined infections determines size. Each triangle represents a single stool sample shown according to the t-distributed stochastic neighbor embedding (t-SNE) algorithm. The axes have arbitrary units with more similar samples clustering together. In upper figure, color corresponds to most abundant taxonomy with filled in triangles signifying that the most abundant taxonomy was dominant (above 90th percentile for age matched controls). In lower figures, color corresponds to either bacterial load (low is below 20th percentile for age matched controls), quantitative phylogenetic diversity, butyrate producers (high above median for age matched controls) or participants age.

#### 2.3 Cox Regression Analysis- Indices, Ratios and Infection Outcomes

| Prior to HCT variable | Bacterial Infections | | | Viral Infections | | | Viral Enterocolitis | | | CDI | | | Bacteremia | | |
|---|---|---|---|---|---|---|---|---|---|---|---|---|---|---|---|
| | HR | 95% CI | P value | HR | 95% CI | P value | HR | 95% CI | P value | HR | 95% CI | P value | HR | 95% CI | P value |
| Bacterial Load (categorical 20th percentile) | 1.03 | [0.54;1.97] | 0.917 | 0.69 | [0.38;1.28] | 0.239 | 0.67 | [0.34;1.36] | 0.27 | 0.88 | [0.39;1.98] | 0.762 | 1.27 | [0.54;2.97] | 0.588 |
| Butyrate Producers (categorical 50th percentile) | 1.27 | [0.55;2.93] | 0.580 | 0.74 | [0.34;1.61] | 0.454 | 0.59 | [0.24;1.50] | 0.269 | 2.88 | [0.81;10.21] | 0.101 | 1.01 | [0.31;3.29] | 0.991 |
| Phylogenetic Diversity (categorical 20th percentile) | 0.84 | [0.37;1.94] | 0.686 | 0.93 | [0.43;2.02] | 0.85 | 1.21 | [0.50;2.96] | 0.676 | 0.49 | [0.17;1.42] | 0.189 | 2.67 | [0.78;9.14] | 0.117 |
| Single Dominant Taxa (categorical 90th percentile) | 0.62 | [0.26;1.45] | 0.271 | 1.22 | [0.57;2.60] | 0.603 | 1.10 | [0.45;2.65] | 0.840 | 0.14 | [0.03;0.64] | 0.011 | 1.29 | [0.44;3.80] | 0.650 |
| ratio 9.0 | 1 | [0.79;1.26] | 0.974 | 1.16 | [0.93;1.44] | 0.186 | 1.09 | [0.85;1.39] | 0.512 | 1.01 | [0.74;1.37] | 0.965 | 0.97 | [0.70;1.33] | 0.849 |
| ratio 12.0 | 0.94 | [0.76;1.16] | 0.574 | 1.24 | [1.04;1.49] | 0.018 | 1.04 | [0.83;1.30] | 0.756 | 0.82 | [0.62;1.09] | 0.164 | 1.15 | [0.87;1.53] | 0.314 |
| ratio 18.0 | 1.03 | [0.84;1.25] | 0.806 | 1.72 | [1.32;2.23] | <0.001 | 1.32 | [1.03;1.70] | 0.029 | 1.03 | [0.79;1.36] | 0.816 | 0.95 | [0.73;1.23] | 0.68 |
| ratio 5.0 | 0.77 | [0.59;0.99] | 0.044 | 1.01 | [0.81;1.27] | 0.911 | 1.18 | [0.91;1.53] | 0.209 | 0.67 | [0.47;0.96] | 0.029 | 0.97 | [0.71;1.34] | 0.867 |
| ratio 11.26 | 1.72 | [1.25;2.37] | <0.001 | 1.39 | [1.04;1.85] | 0.025 | 1.28 | [0.94;1.73] | 0.113 | 1.65 | [1.13;2.41] | 0.009 | 1.24 | [0.86;1.80] | 0.251 |
| ratio 16.16 | 1.11 | [0.86;1.44] | 0.426 | 1.66 | [1.24;2.23] | <0.001 | 2.76 | [1.75;4.36] | <0.001 | 1.23 | [0.89;1.70] | 0.209 | 1.08 | [0.77;1.52] | 0.647 |
| ratio 25.26 | 1.51 | [1.12;2.04] | 0.007 | 1.27 | [0.99;1.62] | 0.063 | 1.14 | [0.85;1.52] | 0.376 | 1.65 | [1.13;2.42] | 0.01 | 1.01 | [0.70;1.46] | 0.969 |
| ratio 3.3 | 1.06 | [0.78;1.44] | 0.702 | 1.35 | [0.99;1.86] | 0.061 | 1.13 | [0.79;1.61] | 0.503 | 1.41 | [0.95;2.09] | 0.085 | 0.6 | [0.36;1.00] | 0.048 |
| ratio 3.16 | 1.23 | [1.00;1.51] | 0.055 | 1.21 | [1.00;1.46] | 0.056 | 1.09 | [0.87;1.37] | 0.464 | 1.1 | [0.87;1.40] | 0.43 | 1.18 | [0.87;1.61] | 0.285 |
| ratio 7.8 | 1.45 | [1.11;1.88] | 0.006 | 1.09 | [0.86;1.38] | 0.484 | 0.98 | [0.75;1.27] | 0.854 | 1.04 | [0.78;1.39] | 0.772 | 3.89 | [2.10;7.21] | <0.001 |

Table ST3: Predictive value of microbiome community indices and compositional ratios of pediatric participants prior to HCT for infectious outcomes. Multiple cox regression model, adjusted for age, gender, conditioning regimen, is displayed as hazard ratio (HR) with 95% confidence interval (CI). P value is adjusted for multiple comparisons.

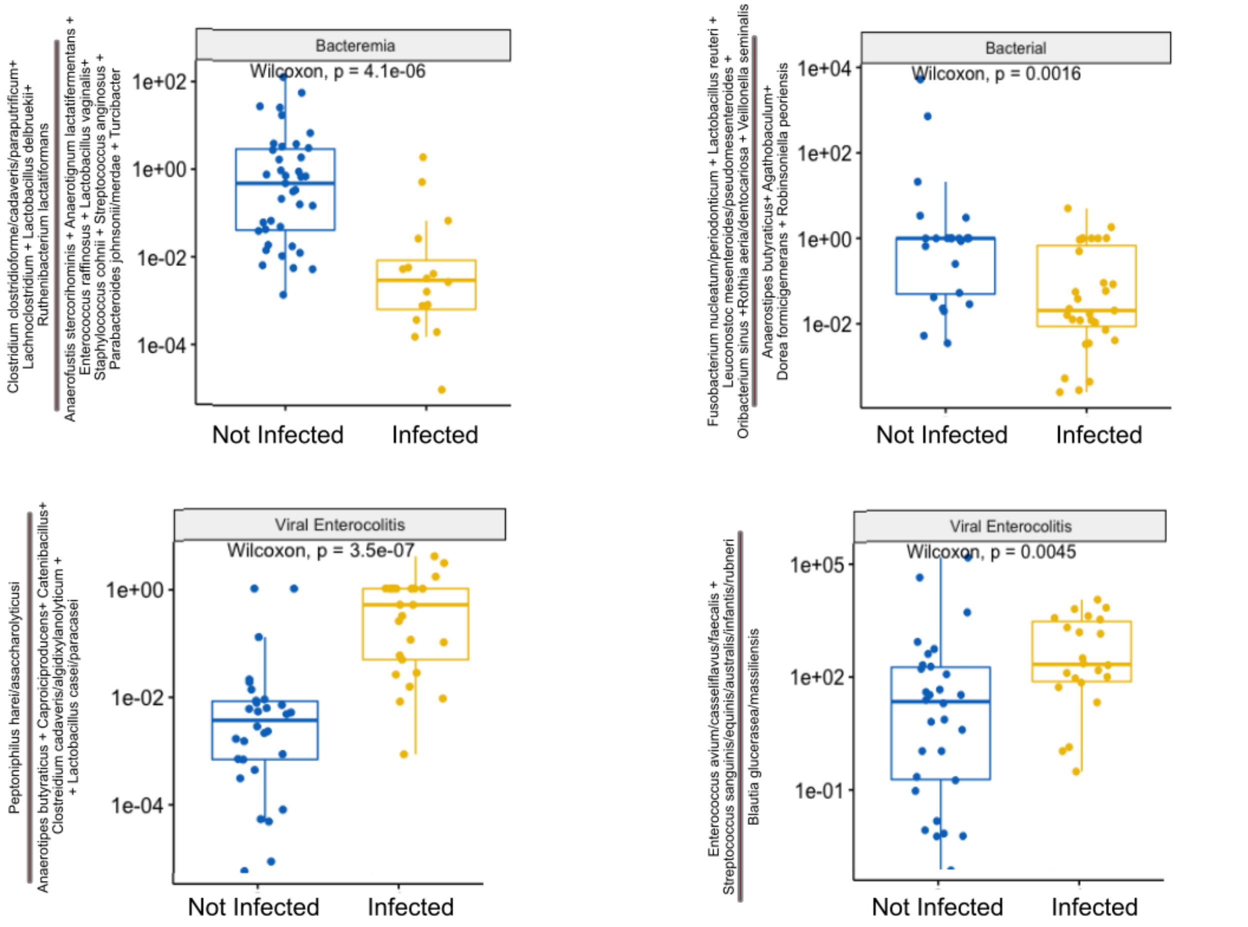

Figure SF8: Four ratios of bacteria species found in the microbiota prior HCT which differed in pediatric HCT participants who have specific infections within year after transplant then those that do not. The phylotypes chosen for these ratios showed similar patterns of differing in abundance and distribution in different communities. Bacterial infections included bacteremias, pneumonias, UTIs and CDI. Viral infections included reactivation of systemic viruses, viral enterocolitis and viral LRTIs. P value is for Wilcoxon Signed Rank Test.

| Neutrophil Recovery variable | Bacterial Infections | | | Viral Infections | | | Viral Enterocolitis | | | CDI | | | Bacteremia | | |
|---|---|---|---|---|---|---|---|---|---|---|---|---|---|---|---|
| | HR | 95% CI | P value | HR | 95% CI | P value | HR | 95% CI | P value | HR | 95% CI | P value | HR | 95% CI | P value |
| Bacterial Load (categorical 20th percentile) | 1.32 | [0.49;3.57] | 0.588 | 1.73 | [0.82;3.64] | 0.15 | 1.22 | [0.49;3.03] | 0.667 | 0.29 | [0.03;2.44] | 0.254 | 1.17 | [0.34;4.03] | 0.803 |
| Butyrate Producers (categorical 50th percentile) | 0.72 | [0.19;2.72] | 0.630 | 0.35 | [0.10;1.28] | 0.113 | 0.35 | [0.07;1.63] | 0.180 | 0.63 | [0.07;6.12] | 0.691 | 0.83 | [0.16;4.20] | 0.819 |
| Phylogenetic Diversity (categorical 20th percentile) | 0.87 | [0.28;2.69] | 0.814 | 2.13 | [0.85;5.33] | 0.107 | 2.41 | [0.78;7.45] | 0.127 | 0.98 | [0.15;6.18] | 0.982 | 0.69 | [0.15;3.17] | 0.632 |
| Single Dominant Taxa (categorical 90th percentile) | 1.24 | [0.38;4.00] | 0.724 | 1.18 | [0.49;2.87] | 0.713 | 0.82 | [0.30;2.25] | 0.699 | 0.79 | [0.14;4.42] | 0.760 | 0.51 | [0.11;2.42] | 0.396 |
| ratio 9.0 | 1.2 | [0.97;1.48] | 0.096 | 1.33 | [1.09;1.62] | 0.006 | 1.35 | [1.05;1.75] | 0.019 | 0.96 | [0.65;1.42] | 0.843 | 1.2 | [0.90;1.60] | 0.224 |
| ratio 12.0 | 1.68 | [1.11;2.55] | 0.015 | 1.25 | [0.92;1.69] | 0.155 | 1.37 | [0.93;2.03] | 0.109 | 1.01 | [0.56;1.82] | 0.974 | 1.58 | [0.89;2.82] | 0.12 |
| ratio 18.0 | 0.99 | [0.70;1.39] | 0.947 | 1.15 | [0.85;1.56] | 0.357 | 1.12 | [0.80;1.58] | 0.505 | 0.98 | [0.55;1.75] | 0.936 | 0.77 | [0.48;1.25] | 0.298 |
| ratio 5.0 | 1.44 | [1.09;1.89] | 0.01 | 1.02 | [0.81;1.29] | 0.861 | 1.09 | [0.84;1.42] | 0.523 | 1.17 | [0.75;1.83] | 0.481 | 2.25 | [1.33;3.78] | 0.002 |
| ratio 11.26 | 0.89 | [0.57;1.39] | 0.601 | 0.99 | [0.72;1.34] | 0.925 | 1.01 | [0.70;1.46] | 0.972 | 0.91 | [0.47;1.76] | 0.777 | 0.69 | [0.36;1.31] | 0.26 |
| ratio 16.16 | 1.34 | [0.87;2.07] | 0.188 | 1 | [0.74;1.36] | 0.986 | 1.23 | [0.85;1.77] | 0.279 | 1.6 | [0.76;3.38] | 0.218 | 1.77 | [0.87;3.59] | 0.116 |
| ratio 25.26 | 0.71 | [0.43;1.18] | 0.189 | 1.04 | [0.73;1.49] | 0.829 | 0.94 | [0.60;1.47] | 0.785 | 0.56 | [0.25;1.25] | 0.16 | 0.59 | [0.30;1.15] | 0.119 |
| ratio 3.3 | 1.49 | [0.99;2.23] | 0.055 | 1.09 | [0.80;1.50] | 0.586 | 1.48 | [1.01;2.18] | 0.045 | 2.48 | [1.35;4.58] | 0.004 | 1.01 | [0.59;1.73] | 0.966 |
| ratio 3.16 | 1.81 | [1.30;2.52] | <0.001 | 1.47 | [1.11;1.94] | 0.007 | 1.96 | [1.38;2.79] | <0.001 | 1.73 | [1.06;2.83] | 0.028 | 1.52 | [0.93;2.48] | 0.096 |
| ratio 7.8 | 0.91 | [0.66;1.25] | 0.56 | 1.12 | [0.88;1.42] | 0.349 | 0.88 | [0.65;1.20] | 0.43 | 0.78 | [0.51;1.22] | 0.277 | 1.1 | [0.73;1.68] | 0.64 |

Table ST4: Predictive value of microbiome community indices and compositional ratios of sample at neutrophil recovery for infectious outcomes in pediatric HCT participants. Multiple cox regression model, adjusted for age, gender, conditioning regimen, is displayed as hazard ratio (HR) with 95% confidence interval (CI). P value is adjusted for multiple comparisons.

None of the phylotype ratios that were predictive of infection in the baseline (prior to HCT) microbiomes were predictive in the neutrophil recovery microbiomes. In fact the only ratio that was significant in both (ratio 5.0) was protective for bacterial infections at baseline and predictive of bacterial infections at neutrophil recovery.

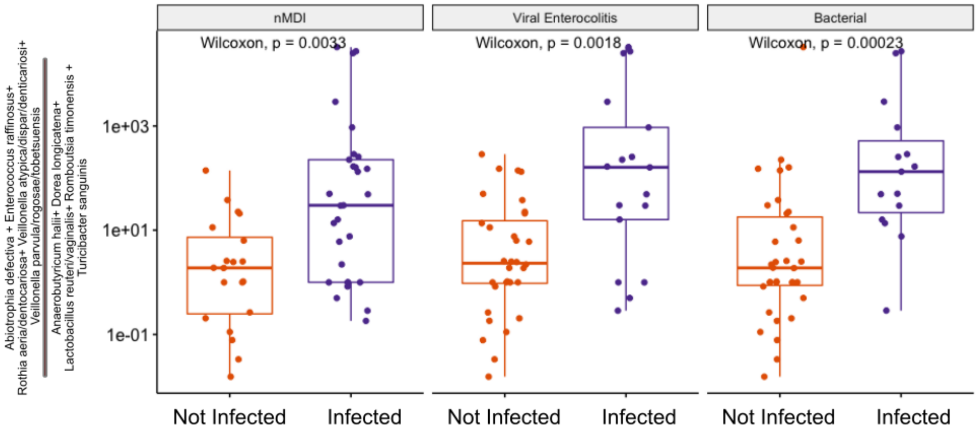

Figure SF9: A single ratio of bacteria species found in the microbiota at time of neutrophil recovery which differed in pediatric HCT participants who had infections or not. The phylotypes chosen for this ratio showed similar patterns of differing in abundance and distribution in different communities. Microbiologically defined infection (nMDI) included any bacterial, viral or parasitic infection. Bacterial infections included bacteremias, pneumonias, UTIs and CDI. P value is for Wilcoxon Signed Rank Test.

#### 2.4 Cox Regression Analysis- Indices, Ratios and Acute GVHD Outcomes

| microbiome variable | Prior to HCT | | | Neutrophil Recovery | | |
|---|---|---|---|---|---|---|
| | HR | 95% CI | P value | HR | 95% CI | P value |
| Bacterial Load (categorical 20th percentile) | 0.94 | [0.43;2.08] | 0.883 | 1.25 | [0.55;2.88] | 0.591 |
| Butyrate Producers (categorical 50 percentile) | 0.95 | [0.35;2.54] | 0.919 | 1.36 | [0.46;4.00] | 0.574 |
| Phylogenetic Diversity (categorical 20th percentile) | 0.47 | [0.15;1.43] | 0.184 | 2.38 | [0.71;7.96] | 0.158 |
| Single Dominant Taxa (categorical 90th percentile) | 0.72 | [0.25;2.08] | 0.541 | 1.01 | [0.38;2.71] | 0.987 |
| ratio 9.0 | 1.65 | [1.22;2.23] | 0.001 | 1.26 | [0.99;1.60] | 0.064 |
| ratio 12.0 | 1.23 | [0.96;1.58] | 0.107 | 1.61 | [1.21;2.16] | 0.001 |
| ratio 18.0 | 1.19 | [0.92;1.54] | 0.19 | 0.94 | [0.71;1.25] | 0.693 |
| ratio 5.0 | 0.93 | [0.70;1.24] | 0.638 | 1.15 | [0.88;1.50] | 0.301 |
| ratio 11.26 | 0.94 | [0.70;1.25] | 0.664 | 1.02 | [0.70;1.49] | 0.919 |
| ratio 16.16 | 1.11 | [0.82;1.52] | 0.497 | 0.94 | [0.69;1.30] | 0.727 |
| ratio 25.26 | 0.84 | [0.62;1.14] | 0.272 | 1.01 | [0.68;1.51] | 0.946 |
| ratio 3.3 | 0.97 | [0.64;1.45] | 0.87 | 0.94 | [0.67;1.30] | 0.689 |
| ratio 3.16 | 0.98 | [0.76;1.27] | 0.879 | 1.3 | [0.94;1.81] | 0.113 |
| ratio 7.8 | 1.45 | [1.02;2.05] | 0.036 | 1.02 | [0.78;1.32] | 0.905 |

Table ST5: Predictive value of microbiome community indices and compositional ratios of samples prior to HCT and at neutrophil recovery for acute GVHD (any grade) in pediatric HCT participants. Multiple cox regression model, adjusted for age, gender, graft type and ex vivo T cell depletion of graft, is displayed as hazard ratio (HR) with 95% confidence interval (CI). P value is adjusted for multiple comparisons.

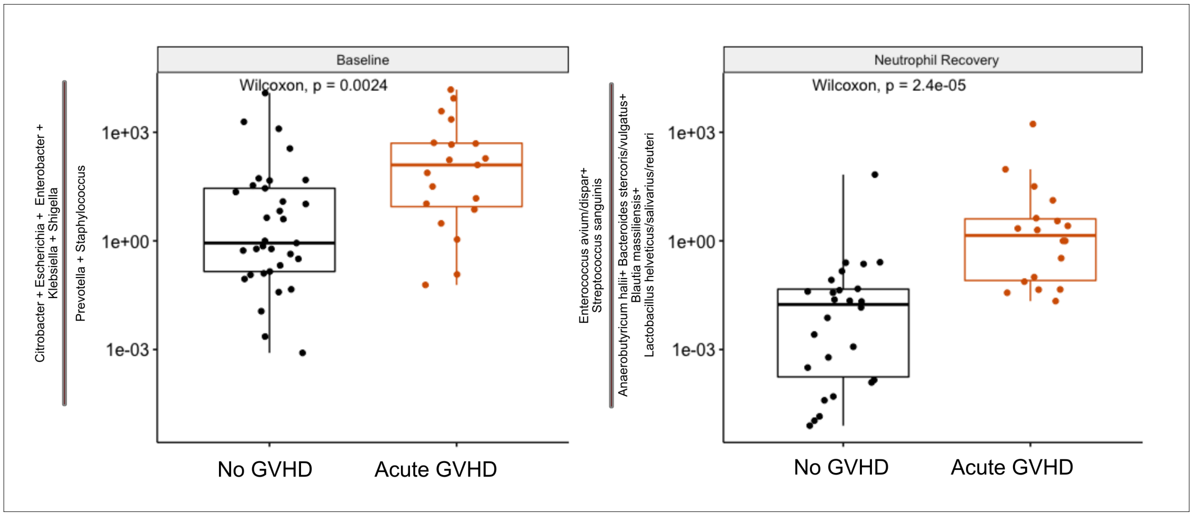

Figure SF10: Two ratios of bacteria species found in the microbiota prior HCT or at neutrophil recovery which differed in pediatric HCT participants who have acute GVHD (any grade) within year after transplant. The phylotypes chosen for these ratios showed similar patterns of differing in abundance and distribution in different communities. P value is for Wilcoxon Signed Rank Test.

#### 3 Phylotypes Included in Ratios

Below is a table of representative sequences from phylotypes which where used in constructing the phylotype ratios.

| V3V4 Sequence | Phylotype | Classified Taxonomy | NCBI |
|---|---|---|---|
| TGGGGAATATTGCACAATGGGGGAAACCCTGATGCAGCGACGCCGCGTGAAGGAAGAAGTATCTCGGTATGTAAACTTCTATCAGCAGGGAAGAAAATGACGGTACCTGACTAAGAAGCCCCGGCTAACTACGTGCCAGCAGCCGCGGTAATACGTAGGGGGCAAGCGTTATCCGGATTTACTGGGTGTAAAGGGAGCGTAGACGGAATAGCAAGTCTGATGTGAAAGGCTGGGGCTTAACCCCAGGACTGCATTGGAAACTGTTGTTCTAGAGTGCCGGAGAGGTAAGCGGAATTCCTAGTGTAGCGGTGAAATGCGTAGATATTAGGAGGAACACCAGTGGCGAAGGCGGCTTACTGGACGGTAACTGACGTTGAGGCTCGAAAGCGTGGGGAGCAAACAGG | pt___00001 | Blautia coccoides or Blautia hansenii | 1532 or 1322 |
| TGGGGAATATTGCACAATGGGGGAAACCCTGATGCAGCGACGCCGCGTGAAGGAAGAAGTATCTCGGTATGTAAACTTCTATCAGCAGGGAAGAAAATGACGGTACCTGACTAAGAAGCCCCGGCTAACTACGTGCCAGCAGCCGCGGTAATACGTAGGGGGCAAGCGTTATCCGGATTTACTGGGTGTAAAGGGAGCGTAGACGGAAGAGCAAGTCTGATGTGAAAGGCAGGGGCCCAACCCCTGGACTGCATTGGAAACTGTCCTTCTTGAGTGCCGGAGGGGTAAGCGGAATTCCTAGTGTAGCGGTGAAATGCGTAGATATTAGGAGGAACACCAGTGGCGAAGGCGGCTTACTGGACGGTAACTGACGTTGAGGCTCGAAAGCGTGGGGAGCAAACAGG | pt___00001 | Blautia stercoris or Blautia hansenii or Blautia coccoides | 871664 or 1322 or 1532 |
| TGGGGAATATTGCACAATGGGGGAAACCCTGATGCAGCGACGCCGCGTGAAGGAAGAAGTATCTCGGTATGTAAACTTCTATCAGCAGGGAAGAAAATGACGGTACCTGACTAAGAAGCCCCGGCTAACTACGTGCCAGCAGCCGCGGTAATACGTAGGGGGCAAGCGTTATCCGGATTTACTGGGTGTAAAGGGAGCGTAGACGGAGCAGCAAGTCTGATGTGAAAGGCGGGGGCTCAACCCCCGGACTGCATTGGAAACTGTTGGTCTTGAGTACCGGAGAGGTAAGCGGAATTCCTAGTGTAGCGGTGAAATGCGTAGATATTAGGAGGAACACCAGTGGCGAAGGCGGCTTACTGGACGGTAACTGACGTTGAGGCTCGAAAGCGTGGGGAGCAAACAGG | pt___00001 | Blautia stercoris or Blautia coccoides or Blautia hansenii | 871664 or 1532 or 1322 |
| TGGGGAATCTTCCGCAATGGACGAAAGTCTGACGGAGCAACGCCGCGTGAGTGATGACGGCCTTCGGGTTGTAAAGCTCTGTTAATCGGGACGAATGGTTCTTGTGCGAATAGTGCGAGGATTTGACGGTACCGGAATAGAAAGCCACGGCTAACTACGTGCCAGCAGCCGCGGTAATACGTAGGTGGCAAGCGTTGTCCGGAATTATTGGGCGTAAAGCGCGCGCAGGCGGATCAGTTAGTCTGTCTTAAAAGTTCGGGGCTTAACCCCGTGATGGGATGGAAACTGCTGATCTAGAGTATCGGAGAGGAAAGTGGAATTCCTAGTGTAGCGGTGAAATGCGTAGATATTAGGAAGAACACCAGTGGCGAAGGCGACTTTCTGGACGAAAACTGACGCTGAGGCGCGAAAGCCAGGGGAGCGAACGGG | pt___00002 | Veillonella atypica | 39777 |
| TGGGGAATCTTCCGCAATGGACGAAAGTCTGACGGAGCAACGCCGCGTGAGTGATGACGGCCTTCGGGTTGTAAAGCTCTGTTAATCGGGACGAAAGGCCTTCTTACGAATAGTTAGAAGGATTGACGGTACCGGAATAGAAAGCCACGGCTAACTACGTGCCAGCAGCCGCGGTAATACGTAGGTGGCAAGCGTTGTCCGGAATTATTGGGCGTAAAGCGCGCGCAGGCGGATTGGTCAGTCTGTCTTAAAAGTTCGGGGCTTAACCCCGTGATGGGATGGAAACTGCCAATCTAGAGTATCGGAGAGGAAAGTGGAATTCCTAGTGTAGCGGTGAAATGCGTAGATATTAGGAAGAACACCAGTGGCGAAGGCGACTTTCTGGACGAAAACTGACGCTGAGGCGCGAAAGCCAGGGGAGCGAACGGG | pt___00002 | Veillonella dispar | 39778 |
| TGGGGAATATTGCACAATGGGCGGAAGCCTGATGCAGCGACGCCGCGTGAGGGATGGAGGCCTTCGGGTTGTGAACCTCTTTCGCCCGTGGTCAAGCCGCAACTGTGGGTTGTGGTGAGGGTAGTGGGTAAAGAAGCGCCGGCTAACTACGTGCCAGCAGCCGCGGTAATACGTAGGGCGCGAGCGTTGTCCGGAATTATTGGGCGTAAAGGGCTTGTAGGCGGCTGGTCGCGTCTGCCGTGAAATCCTCTGGCTCAACTGGGGGCGTGCGGTGGGTACGGGCTGGCTTGAGTGCGGTAGGGGAGGCTGGAACTCCTGGTGTAGCGGTGGAATGCGCAGATATCAGGAAGAACACCGGTGGCGAAGGCGGGTCTCTGGGCCGTTACTGACGCTGAGGAGCGAAAGCGTGGGGAGCGAACAGG | pt___00003 | Schaalia georgiae | 52768 |
| TGGGGAATATTGCACAATGGGCGAAAGCCTGATGCAGCAACGCCGCGTGAGCGATGAAGGCCTTCGGGTCGTAAAGCTCTGTCCTCAAGGAAGATAATGACGGTACTTGAGGAGGAAGCCCCGGCTAACTACGTGCCAGCAGCCGCGGTAATACGTAGGGGGCTAGCGTTATCCGGAATTACTGGGCGTAAAGGGTGCGTAGGTGGTTTCTTAAGTCAGAAGTGAAAGGCTACGGCTCAACCGTAGTAAGCTTTTGAAACTGAGAAACTTGAGTGCAGGAGAGGAGCGTAGAATTCCTAGTGTAGCGGTGAAATGCGTAGATATTAGGAGGAATACCAGTTGCGAAGGCGGCTCTCTGGACTGTAACTGACACTGAGGCACGAAAGCGTGGGGAGCAAACAGG | pt___00005 | Romboutsia sedimentorum | 1368474 |
| TGGGGAATATTGCACAATGGGCGCAAGCCTGATGCAGCCATGCCGCGTGTATGAAGAAGGCCTTCGGGTTGTAAAGTACTTTCAGCGAGGAGGAAGGCATTAAGGTTAATAACCTTAGTGATTGACGTTACTCGCAGAAGAAGCACCGGCTAACTCCGTGCCAGCAGCCGCGGTAATACGGAGGGTGCAAGCGTTAATCGGAATTACTGGGCGTAAAGCGCACGCAGGCGGTCTGTTAAGTCAGATGTGAAATCCCCGGGCTCAACCTGGGAACTGCATTTGAAACTGGCAGGCTTGAGTCTTGTAGAGGGGGGTAGAATTCCAGGTGTAGCGGTGAAATGCGTAGAGATCTGGAGGAATACCGGTGGCGAAGGCGGCCCCCTGGACAAAGACTGACGCTCAGGTGCGAAAGCGTGGGGAGCAAACAGG | pt___00005 | Raoultella ornithinolytica | 54291 |
| TGGGGAATATTGCACAATGGGCGCAAGCCTGATGCAGCCATGCCGCGTGTATGAAGAAGGCCTTCGGGTTGTAAAGTACTTTCAGCGGGGAGGAAGGGAGTGAAGTTAATAACCTCATTCATTGACGTTACCCGCAGAAGAAGCACCGGCTAACTCCGTGCCAGCAGCCGCGGTAATACGGAGGGTGCAAGCGTTAATCGGAATTACTGGGCGTAAAGCGCACGCAGGCGGTCTGTCAAGTCGGATGTGAAATCCCCGGGCTCAACCTGGGAACTGCATTCGAAACTGGCAGGCTGGAGTCTTGTAGAGGGGGGTAGAATTCCAGGTGTAGCGGTGAAATGCGTAGAGATCTGGAGGAATACCGGTGGCGAAGGCGGCCCCCTGGACAAAGACTGACGCTCAGGTGCGAAAGCGTGGGGAGCAAACAGG | pt___00005 | Klebsiella oxytoca | 571 |

| | | | |
|---|---|---|---|
| TGGGGAATATTGCACAATGGGCGCAAGCCTGATGCAGCCATGCCGCGTGTGTGAAGAAGGCCTTCGGGTTGTAAAGCACTT<br>TCAGCGGGGAGGAAGGCGTTAAGGTTAATAACCTTGGCGATTGACGTTACCCGCAGAAGAAGCACCGGCTAACTCCGTGCC<br>AGCAGCCGCGGTAATACGGAGGGTGCAAGCGTTAATCGGAATTACTGGGCGTAAAGCGCACGCAGGCGGTCTGTCAAGTCG<br>GATGTGAAATCCCCGGGCTTAACCTGGGAACTGCATTCGAAACTGGCAGGCTAGAGTCTTGTAGAGGGGGGTAGAATTCCA<br>GGTGTAGCGGTGAAATGCGTAGAGATCTGGAGGAATACCGGTGGCGAAGGCGGCCCCCTGGACAAAGACTGACGCTCAGGT<br>GCGAAAGCGTGGGGAGCAAACAGG | pt___00005 | Klebsiella pneumoniae | 573 |
| TAGGGAATCTTCGGCAATGGGGGGAACCCTGACCGAGCAACGCCGCGTGAGTGAAGAAGGTTTTCGGATCGTAAAGCTCTG<br>TTGTAAGAGAAGAACGGGTGTGAGAGTGGAAAGTTCACACTGTGACGGTATCTTACCAGAAAGGGACGGCTAACTACGTGC<br>CAGCAGCCGCGGTAATACGTAGGTCCCGAGCGTTGTCCGGATTTATTGGGCGTAAAGCGAGCGCAGGCGGTTAGATAAGTC<br>TGAAGTTAAAGGCTGTGGCTTAACCATAGTATGCTTTGGAAACTGTTTAACTTGAGTGCAGAAGGGGAGAGTGGAATTCCA<br>TGTGTAGCGGTGAAATGCGTAGATATATGGAGGAACACCGGTGGCGAAAGCGGCTCTCTGGTCTGTAACTGACGCTGAGGC<br>TCGAAAGCGTGGGGAGCAAACAGG | pt___00007 | Streptococcus sanguinis | 1305 |
| TAGGGAATCTTCGGCAATGGACGCAAGTCTGACCGAGCAACGCCGCGTGAGTGAAGAAGGTTTTCGGATCGTAAAACTCTG<br>TTGTTAGAGAAGAACAAGGATGAGAGTAGAACGTTCATCCCTTGACGGTATCTAACCAGAAAGCCACGGCTAACTACGTGCC<br>AGCAGCCGCGGTAATACGTAGGTGGCAAGCGTTGTCCGGATTTATTGGGCGTAAAGCGAGCGCAGGCGGTTTCTTAAGTCT<br>GATGTGAAAGCCCCCGGCTCAACCGGGGAGGGTCATTGGAAACTGGGAAACTTGAGTGCAGAAGAGGAGAGTGGAATTCCA<br>TGTGTAGCGGTGAAATGCGTAGATATATGGAGGAACACCAGTGGCGAAGGCGGCTCTCTGGTCTGTAACTGACGCTGAGGC<br>TCGAAAGCGTGGGGAGCAAACAGG | pt___00007 | Enterococcus avium | 33945 |
| TAGGGAATCTTCGGCAATGGACGAAAGTCTGACCGAGCAACGCCGCGTGAGTGAAGAAGGTTTTCGGATCGTAAAACTCTG<br>TTGTTAGAGAAGAACAAGGATGAGAGTAGAATGTTCATCCCTTGACGGTATCTAACCAGAAAGCCACGGCTAACTACGTGCC<br>AGCAGCCGCGGTAATACGTAGGTGGCAAGCGTTGTCCGGATTTATTGGGCGTAAAGCGAGCGCAGGCGGTTTCTTAAGTCT<br>GATGTGAAAGCCCCCGGCTCAACCGGGGAGGGTCATTGGAAACTGGGAAACTTGAGTGCAGAAGAGGAGAGTGGAATTCCA<br>TGTGTAGCGGTGAAATGCGTAGATATATGGAGGAACACCAGTGGCGAAGGCGGCTCTCTGGTCTGTAACTGACGCTGAGGC<br>TCGAAAGCGTGGGGAGCAAACAGG | pt___00007 | Enterococcus raffinosus or Enterococcus avium | 71452 or 33945 |
| TAGGGAATCTTCGGCAATGGACGAAAGTCTGACCGAGCAACGCCGCGTGAGTGAAGAAGGTTTTCGGATCGTAAAACTCTG<br>TTGTTAGAGAAGAACAAGGATGAGAGTAGAACGTTCATCCCTTGACGGTATCTAACCAGAAAGCCACGGCTAACTACGTGCC<br>AGCAGCCGCGGTAATACGTAGGTGGCAAGCGTTGTCCGGATTTATTGGGCGTAAAGCGAGCGCAGGCGGTTTCTTAAGTCT<br>GATGTGAAAGCCCCCGGCTCAACCGGGGAGGGTCATTGGAAACTGGGAGACTTGAGTGCAGAAGAGGAGAGTGGAATTCCA<br>TGTGTAGCGGTGAAATGCGTAGATATATGGAGGAACACCAGTGGCGAAGGCGGCTCTCTGGTCTGTAACTGACGCTGAGGC<br>TCGAAAGCGTGGGGAGCGAACAGG | pt___00009 | Enterococcus gallinarum | 1353 |
| TGGGGAATATTGCACAATGGGCGCAAGCCTGATGCAGCCATGCCGCGTGTGTGAAGAAGGCCTTCGGGTTGTAAAGCACTT<br>TCAGCGGGGAGGAAGGCGGTGAGGTTAATAACCTCACCGATTGACGTTACCCGCAGAAGAAGCACCGGCTAACTCCGTGCC<br>AGCAGCCGCGGTAATACGGAGGGTGCAAGCGTTAATCGGAATTACTGGGCGTAAAGCGCACGCAGGCGGTCTGTCAAGTCG<br>GATGTGAAATCCCCGGGCTCAACCTGGGAACTGCATTCGAAACTGGCAGGCTAGAGTCTTGTAGAGGGGGGTAGAATTCCA<br>GGTGTAGCGGTGAAATGCGTAGAGATCTGGAGGAATACCGGTGGCGAAGGCGGCCCCCTGGACAAAGACTGACGCTCAGGT<br>GCGAAAGCGTGGGGAGCAAACAGG | pt___00009 | Klebsiella variicola | 244366 |
| TGGGGAATATTGCACAATGGGGGAAACCCTGATGCAGCGACGCCGCGTGAGCGAAGAAGTATTTCGGTATGTAAAGCTCTA<br>TCAGCAGGGAAGAAAATGACGGTACCTGACTAAGAAGCCCCGGCTAACTACGTGCCAGCAGCCGCGGTAATACGTAGGGGG<br>CAAGCGTTATCCGGATTTACTGGGTGTAAAGGGAGCGTAGACGGCCAGGCAAGTCTGATGTGAAAGGCAGGGGCTCAACCC<br>CTGGACTGCATTGGAAACTGCCAGGCTGGAGTGCCGGAGAGGTAAGCGGAATTCCTAGTGTAGCGGTGAAATGCGTAGATA<br>TTAGGAGGAACACCAGTGGCGAAGGCGGCTTACTGGACGGTAACTGACGTTGATGCTCGAAAGCGTGGGGAGCAAACAGG | pt___00015 | Murimonas intestini | 1337051 |
| TAGGGAATCTTCGGCAATGGGGGCAACCCTGACCGAGCAACGCCGCGTGAGTGAAGAAGGTTTTCGGATCGTAAAGCTCTG<br>TTGTAAGTCAAGAACGGGTGTGAGAGTGGAAAGTTCACACTATGACGGTAGCTTACCAGAAAGGGACGGCTAACTACGTGC<br>CAGCAGCCGCGGTAATACGTAGGTCCCGAGCGTTGTCCGGATTTATTGGGCGTAAAGCGAGCGCAGGCGGTTTGATAAGTC<br>TGAAGTTAAAGGCTGTGGCTCAACCATAGTTCGCTTTGGAAACTGTCAAACTTGAGTGCAGAAGGGGAGAGTGGAATTCCA<br>TGTGTAGCGGTGAAATGCGTAGATATATGGAGGAACACCGGTGGCGAAAGCGGCTCTCTGGTCTGTAACTGACGCTGAGGC<br>TCGAAAGCGTGGGGAGCGAACAGG | pt___00016 | Streptococcus salivarius | 1304 |
| TGGGGAATATTGCACAATGGGGGAAACCCTGATGCAGCAACGCCGCGTGAGTGATGACGGCCTTCGGGTTGTAAAGCTCTG<br>TCTTCAGGGACGATAATGACGGTACCTGAGAAGGAAGCCACGGCTAACTACGTGCCAGCAGCCGCGGTAATACGTAGGTGG<br>CGAGCGTTGTCCGGATTTACTGGGCGTAAAGGGAGCGTAGGCGGACTTTTAAGTGAGATGTGAAATACCCGGGCTCAACTT<br>GGGTGCTGCATTTCAAACTGGAAGTCTAGAGTGCAGGAGAGGAGAATGGAATTCCTAGTGTAGCGGTGAAATGCGTAGAGA<br>TTAGGAAGAACACCAGTGGCGAAGGCGATTCTCTGGACTGTAACTGACGCTGAGGCTCGAAAGCGTGGGGAGCAAACAGG | pt___00016 | Clostridium disporicum | 84024 |

| | | | |
|---|---|---|---|
| TGGGGAATATTGCACAATGGGCGCAAGCCTGATGCAGCCATGCCGCGTGTATGAAGAAGGCCTTCGGGTTGTAAAGTACTTTCAGCGAGGAGGAAGGCGTTGTGGTTAATAACCGCAACGATTGACGTTACTCGCAGAAGAAGCACCGGCTAACTCCGTGCCAGCAGCCGCGGTAATACGGAGGGTGCAAGCGTTAATCGGAATTACTGGGCGTAAAGCGCACGCAGGCGGTCTGTCAAGTCGGATGTGAAATCCCCGGGCTCAACCTGGGAACTGCATCCGAAACTGGCAGGCTAGAGTCTTGTAGAGGGGGGTAGAATTCCAGGTGTAGCGGTGAAATGCGTAGAGATCTGGAGGAATACCGGTGGCGAAGGCGGCCCCCTGGACAAAGACTGACGCTCAGGTGCGAAAGCGTGGGGAGCAAACAGG | pt___00018 | Citrobacter freundii | 546 |
| TAGGGAATCTTCGGCAATGGACGAAAGTCTGACCGAGCAACGCCGCGTGAGTGAAGAAGGTTTTCGGATCGTAAAACTCTGTTGTTAGAGAAGAACAAGGACGTTAGTAACTGAACGTCCCCTGACGGTATCTAACCAGAAAGCCACGGCTAACTACGTGCCAGCAGCCGCGGTAATACGTAGGTGGCAAGCGTTGTCCGGATTTATTGGGCGTAAAGCGAGCGCAGGCGGTTTCTTAAGTCTGATGTGAAAGCCCCCGGCTCAACCGGGGAGGGTCATTGGAAACTGGGAAACTTGAGTGCAGAAGAGGAGAGTGGAATTCCATGTGTAGCGGTGAAATGCGTAGATATATGGAGGAACACCAGTGGCGAAGGCGGCTCTCTGGTCTGTAACTGACGCTGAGGCTCGAAAGCGTGGGGAGCAAACAGG | pt___00022 | Enterococcus faecalis | 1351 |
| TGAGGAATATTGGTCAATGGGCGATGGCCTGAACCAGCCAAGTAGCGTGAAGGATGACTGCCCTATGGGTTGTAAACTTCTTTTATAAAGGAATAAAGTCGGGTATGCATACCCGTTTGCATGTACTTTATGAATAAGGATCGGCTAACTCCGTGCCAGCAGCCGCGGTAATACGGAGGATCCGAGCGTTATCCGGATTTATTGGGTTTAAAGGGAGCGTAGATGGATGTTTAAGTCAGTTGTGAAAGTTTGCGGCTCAACCGTAAAATTGCAGTTGATACTGGATGTCTTGAGTGCAGTTGAGGCAGGCGGAATTCGTGGTGTAGCGGTGAAATGCTTAGATATCACGAAGAACTCCGATTGCGAAGGCAGCCTGCTAAGCTGCAACTGACATTGAGGCTCGAAAGTGTGGGTATCAAACAGG | pt___00022 | Phocaeicola dorei | 357276 |
| TAGGGAATCTTCGGCAATGGGCGAAAGCCTGACCGAGCAACGCCGCGTGAATGATGAAGGCCTTCGGGTTGTAAAATTCTGTTATAAGGGAAGAATGGCTCTAGTAGGAAATGGCTAGAGTGTGACGGTACCTTATGAGAAAGCCACGGCTAACTACGTGCCAGCAGCCGCGGTAATACGTAGGTGGCGAGCGTTATCCGGAATTATTGGGCGTAAAGAGCGCGCAGGTGGTTGATTAAGTCTGATGTGAAAGCCCACGGCTTAACCGTGGAGGGTCATTGGAAACTGGTCAACTTGAGTGCAGAAGAGGGAAGTGGAATTCCATGTGTAGCGGTGAAATGCGTAGAGATATGGAGGAACACCAGTGGCGAAGGCGGCTTCCTGGTCTGTAACTGACACTGAGGCGCGAAAGCGTGGGGAGCAAACAGG | pt___00023 | Turicibacter sanguinis | 154288 |
| TGGGGAATATTGCACAATGGGGGAAACCCTGATGCAGCAACGCCGCGTGAGTGATGACGGTCTTCGGATTGTAAAGCTCTGTCTTTAGGGACGATAATGACGGTACCTAAGGAGGAAGCCACGGCTAACTACGTGCCAGCAGCCGCGGTAATACGTAGGTGGCAAGCGTTGTCCGGATTTACTGGGCGTAAAGGGAGCGTAGGTGGATATTTAAGTGGGATGTGAAATACCCGGGCTTAACCTGGGTGCTGCATTCCAAACTGGATATCTAGAGTGCAGGAGAGGAAAGGAGAATTCCTAGTGTAGCGGTGAAATGCGTAGAGATTAGGAAGAATACCAGTAGCGAAGGCGCCTTTCTGGACTGTAACTGACACTGAGGCTCGAAAGCGTGGGGAGCAAACAGG | pt___00025 | Clostridium butyricum | 1492 |
| TGGGGAATATTGCACAATGGGCGCAAGCCTGATGCAGCGACGCCGCGTGAGTGAAGAAGTATCTCGGTATGTAAAGCTCTATCAGCAGGGAAGAAAATGACGGTACCTGACTAAGAAGCCCCGGCTAACTACGTGCCAGCAGCCGCGGTAATACGTAGGGGGCAAGCGTTATCCGGATTTACTGGGTGTAAAGGGAGCGTAGACGGCGACGCAAGTCTGGAGTGAAAGCCCGGGGCCCAACCCCGGGACTGCTTTGGAAACTGTGCTGCTGGAGTGCAGGAGAGGTAAGTGGAATTCCTAGTGTAGCGGTGAAATGCGTAGATATTAGGAGGAACACCAGTGGCGAAGGCGGCTTACTGGACTGTAACTGACGTTGAGGCTCGAAAGCGTGGGGAGCAAACAGG | pt___00025 | Enterocloster aldenensis | 358742 |
| TAGGGAATCTTCCGCAATGGGCGAAAGCCTGACGGAGCAACGCCGCGTGAGTGATGAAGGTCTTCGGATCGTAAAACTCTGTTATCAGGGAAGAACAAACGTGTAAGTAACTGTGCACGTCTTGACGGTACCTGATCAGAAAGCCACGGCTAACTACGTGCCAGCAGCCGCGGTAATACGTAGGTGGCAAGCGTTATCCGGAATTATTGGGCGTAAAGCGCGCGTAGGCGGTTTTTTAAGTCTGATGTGAAAGCCCACGGCTCAACCGTGGAGGGTCATTGGAAACTGGAAAACTTGAGTGCAGAAGAGGAAAGTGGAATTCCATGTGTAGCGGTGAAATGCGCAGAGATATGGAGGAACACCAGTGGCGAAGGCGACTTTCTGGTCTGTAACTGACGCTGATGTGCGAAAGCGTGGGGATCAAACAGG | pt___00027 | Staphylococcus pasteuri | 45972 |
| TAGGGAATCTTCGGCAATGGGGGCAACCCTGACCGAGCAACGCCGCGTGAGTGAAGAAGGTTTTCGGATCGTAAAGCTCTGTTGTAAGTCAAGAACGGGTGTGAGAGTGGAAAGTTCACACTGTGACGGTAGCTTACCAGAAAGGGACGGCTAACTACGTGCCAGCAGCCGCGGTAATACGTAGGTCCCGAGCGTTGTCCGGATTTATTGGGCGTAAAGCGAGCGCAGGCGGTTTGATAAGTCTGAAGTTAAAGACTGTGGCTCAACCATAGTTCGCTTTGGAAACTGTCAAACTTGAGTGCAGAAGGGGAGAGTGGAATTCCATGTGTAGCGGTGAAATGCGTAGATATATGGAGGAACACCGGTGGCGAAAGCGGCTCTCTGGTCTGTAACTGACGCTGAGGCTCGAAAGCGTGGGGAGCGAACAGG | pt___00031 | Streptococcus salivarius or Streptococcus vestibularis | 1304 or 1343 |
| TAGGGAATCTTCGGCAATGGGGGCAACCCTGACCGAGCAACGCCGCGTGAGTGAAGAAGGTTTTCGGATCGTAAAGCTCTGTTGTAAGAGAAGAACGAGTGTGAGAGTGGAAAGTTCACACTGTGACGGTATCTTACCAGAAAGGGACGGCTAACTACGTGCCAGCAGCCGCGGTAATACGTAGGTCCCGAGCGTTGTCCGGATTTATTGGGCGTAAAGCGAGCGCAGGCGGTTAGATAAGTCTGAAGTTAAAGGCTGTGGCTTAACCATAGTACGCTTTGGAAACTGTTTAACTTGAGTGCAGAAGGGGAGAGTGGAATTCCATGTGTAGCGGTGAAATGCGTAGATATATGGAGGAACACCGGTGGCGAAAGCGGCTCTCTGGTCTGTAACTGACGCTGAGGCTCGAAAGCGTGGGGAGCAAACAGG | pt___00033 | Streptococcus | 1301 |

| | | | |
|---|---|---|---|
| TGGGGAATATTGCACAATGGGCGCAAGCCTGATGCAGCGACGCCGCGTGAGGGATGGAGGCCTTCGGGTTGTAAACCTCTT<br>TTGTTAGGGAGCAAGGCACTTTGTGTTGAGTGTACCTTTCGAATAAGCACCGGCTAACTACGTGCCAGCAGCCGCGGTAATA<br>CGTAGGGTGCAAGCGTTATCCGGAATTATTGGGCGTAAAGGGCTCGTAGGCGGTTCGTCGCGTCCGGTGTGAAAGTCCATC<br>GCTTAACGGTGGATCCGCGCCGGGTACGGGCGGGCTTGAGTGCGGTAGGGGAGACTGGAATTCCCGGTGTAACGGTGGAAT<br>GTGTAGATATCGGGAAGAACACCAATGGCGAAGGCAGGTCTCTGGGCCGTTACTGACGCTGAGGAGCGAAAGCGTGGGGAG<br>CGAACAGG | pt___00039 | Bifidobacterium breve | 1685 |
| TGGGGAATATTGCACAATGGGCGCAAGCCTGATGCAGCGACGCCGCGTGAGGGATGGAGGCCTTCGGGTTGTAAACCTCTT<br>TTATCGGGGAGCAAGCGAGAGTGAGTTTACCCGTTGAATAAGCACCGGCTAACTACGTGCCAGCAGCCGCGGTAATACGTA<br>GGGTGCAAGCGTTATCCGGAATTATTGGGCGTAAAGGGCTCGTAGGCGGTTCGTCGCGTCCGGTGTGAAAGTCCATCGCTT<br>AACGGTGGATCCGCGCCGGGTACGGGCGGGCTTGAGTGCGGTAGGGGAGACTGGAATTCCCGGTGTAACGGTGGAATGTGT<br>AGATATCGGGAAGAACACCAATGGCGAAGGCAGGTCTCTGGGCCGTTACTGACGCTGAGGAGCGAAAGCGTGGGGAGCGAA<br>CAGG | pt___00039 | Bifidobacterium longum | 216816 |
| TAGGGAATCTTCCACAATGGGCGCAAGCCTGATGGAGCAACACCGCGTGAGTGAAGAAGGGTTTCGGCTCGTAAAGCTCTG<br>TTGTTAAAGAAGAACACGTATGAGAGCAACTGTTCATACGTTGACGGTATTTAACCAGAAAGTCACGGCTAACTACGTGCCA<br>GCAGCCGCGGTAATACGTAGGTGGCAAGCGTTATCCGGATTTATTGGGCGTAAAGAGAGTGCAGGCGGTTTTCTAAGTCTG<br>ATGTGAAAGCCTTCGGCTTAACCGGAGAAGTGCATCGGAAACTGGATAACTTGAGTGCAGAAGAGGGTAGTGGAACTCCAT<br>GTGTAGCGGTGGAATGCGTAGATATATGGAAGAACACCAGTGGCGAAGGCGGCTACCTGGTCTGCAACTGACGCTGAGACT<br>CGAAAGCATGGGTAGCGAACAGG | pt___00042 | Limosilactobacillus fermentum | 1613 |
| TGAGGAATATTGGTCAATGGGCGCTAGCCTGAACCAGCCAAGTAGCGTGAAGGATGAAGGCTCTATGGGTCGTAAACTTCT<br>TTTATATAAGAATAAAGTGCAGTATGTATACTGTTTTGTATGTATTATATGAATAAGGATCGGCTAACTCCGTGCCAGCAGC<br>CGCGGTAATACGGAGGATCCGAGCGTTATCCGGATTTATTGGGTTTAAAGGGAGCGTAGGTGGACTGGTAAGTCAGTTGTG<br>AAAGTTTGCGGCTCAACCGTAAAATTGCAGTTGAAACTGGCAGTCTTGAGTACAGTAGAGGTGGGCGGAATTCGTGGTGTA<br>GCGGTGAAATGCTTAGATATCACGAAGAACTCCGATTGCGAAGGCAGCTCACTAGACTGCAACTGACACTGATGCTCGAAAG<br>TGTGGGTATCAAACAGG | pt___00042 | Bacteroides fragilis | 817 |
| TGGGGAATATTGCACAATGGGCGCAAGCCTGATGCAGCGACGCCGCGTGAGGGATGGAGGCCTTCGGGTTGTGAACCTCTT<br>TCGCCAGTGAAGCAGGCCTGCCTCGTTTGTGGGTGGGTTGACGGTAGCTGGATAAGAAGCGCCGGCTAACTACGTGCCAGC<br>AGCCGCGGTAATACGTAGGGCGCGAGCGTTGTCCGGAATTATTGGGCGTAAAGAGCTCGTAGGCGGCTGGTCGCGTCTGTC<br>GTGAAATCCTCTGGCTTAACTGGGGGCTTGCGGTGGGTACGGGCCGGCTTGAGTGCGGTAGGGGAGACTGGAACTCCTGGT<br>GTAGCGGTGGAATGCGCAGATATCAGGAAGAACACCGGTGGCGAAGGCGGGTCTCTGGGCCGTTACTGACGCTGAGGAGCG<br>AAAGCGTGGGGAGCGAACAGG | pt___00043 | Actinomyces naeslundii | 1655 |
| TGGGGAATATTGCACAATGGGCGCAAGCCTGATGCAGCGACGCCGCGTGAGGGATGGAGGCCTTCGGGTTGTAAACCTCTT<br>TCGCCAGTGAAGCAGGCCTGCCTCGTTTGTGGGTGGGTTGACGGTAGCTGGATAAGAAGCGCCGGCTAACTACGTGCCAGC<br>AGCCGCGGTAATACGTAGGGCGCGAGCGTTGTCCGGAATTATTGGGCGTAAAGAGCTCGTAGGCGGCTGGTCGCGTCTGTC<br>GTGAAATCCTCTGGCTTAACTGGGGGCTTGCGGTGGGTACGGGCCGGCTTGAGTGCGGTAGGGGAGACTGGAACTCCTGGT<br>GTAGCGGTGGAATGCGCAGATATCAGGAAGAACACCGGTGGCGAAGGCGGGTCTCTGGGCCGTTACTGACGCTGAGGAGCG<br>AAAGCGTGGGGAGCGAACAGG | pt___00043 | Actinomyces naeslundii or Actinomyces oris | 1655 or 544580 |
| TGGGGAATATTGCACAATGGGCGCAAGCCTGATGCAGCGACGCCGCGTGAGGGATGGAGGCCTTCGGGTTGTAAACCTCTT<br>TCGCCAGTGAAGCAGGCCTGTCCCTTTTGTGGGTGGGTTGACGGTAGCTGGATAAGAAGCGCCGGCTAACTACGTGCCAGC<br>AGCCGCGGTAATACGTAGGGCGCGAGCGTTGTCCGGAATTATTGGGCGTAAAGAGCTCGTAGGCGGCTGGTCGCGTCTGTC<br>GTGAAATCCTCTGGCTTAACTGGGGGCTTGCGGTGGGTACGGGCCGGCTTGAGTGCGGTAGGGGAGACTGGAACTCCTGGT<br>GTAGCGGTGGAATGCGCAGATATCAGGAAGAACACCGGTGGCGAAGGCGGGTCTCTGGGCCGTTACTGACGCTGAGGAGCG<br>AAAGCGTGGGGAGCGAACAGG | pt___00043 | Actinomyces oris | 544580 |
| TGAGGAATATTGGTCAATGGGCGAGAGCCTGAACCAGCCAAGTAGCGTGCAGGATGACGGCCCTATGGGTTGTAAACTGCT<br>TTTGTATGGGGATAAAGTCAGTCACGTGTGATTGTTTGCAGGTACCATACGAATAAGGACCGGCTAATTCCGTGCCAGCAGC<br>CGCGGTAATACGGAAGGTCCGGGCGTTATCCGGATTTATTGGGTTTAAAGGGAGCGTAGGCTGGAGATTAAGTGTGTTGTG<br>AAATGTAGACGCTCAACGTCTGACTTGCAGCGCATACTGGTTTCCTTGAGTACGCACAACGTTGGCGGAATTCGTCGTGTAG<br>CGGTGAAATGCTTAGATATGACGAAGAACTCCGATTGCGAAGGCAGCTGACGGGAGCGCAACTGACGCTGAAGCTCGAAGG<br>TGCGGGTATCGAACAGG | pt___00046 | Prevotella histicola | 470565 |
| TGGGGAATATTGCACAATGGGGGAAACCCTGATGCAGCGACGCCGCGTGAGTGAAGAAGTATCTCGGTATGTAAAGCTCTA<br>TCAGCAGGGAAGAAAATGACGGTACCTGACTAAGAAGCCCCGGCTAACTACGTGCCAGCAGCCGCGGTAATACGTAGGGGG<br>CAAGCGTTATCCGGAATTACTGGGTGTAAAGGGTGCGTAGGTGGTATGGCAAGTCAGAAGTGAAAACCCAGGGCTTAACTC<br>TGGGACTGCTTTTGAAACTGTCAGACTGGAGTGCAGGAGAGGTAAGCGGAATTCCTAGTGTAGCGGTGAAATGCGTAGATA<br>TTAGGAGGAACATCAGTGGCGAAGGCGGCTTACTGGACTGAAACTGACACTGAGGCACGAAAGCGTGGGGAGCAAACAGG | pt___00047 | Anaerostipes hadrus | 649756 |

| | | | |
|---|---|---|---|
| TGGGGGATATTGCACAATGGGGGAAACCCTGATGCAGCAACGCCGCGTGAGGGAAGAAGGTTTTCGGATTGTAAACCTCTGTTCTTAGTGACGATAATGACGGTAGCTAAGGAGAAAGCTCCGGCTAACTACGTGCCAGCAGCCGCGGTAATACGTAGGGAGCGAGCGTTGTCCGGATTTACTGGGTGTAAAGGGTGCGTAGGCGGCGAGGCAAGTCAGGCGTGAAATCTATGGGCTTAACCCATAAACTGCGCTTGAAACTGTCTTGCTTGAGTGAAGTAGAGGTAGGCGGAATTCCCGGTGTAGCGGTGAAATGCGTAGAGATCGGGAGGAACACCAGTGGCGAAGGCGGCCTACTGGGCTTTAACTGACGCTGAAGCACGAAAGCATGGGTAGCAAACAGG | pt___00053 | [Clostridium] leptum | 1535 |
| TAGGGAATCTTCGGCAATGGACGAAAGTCTGACCGAGCAACGCCGCGTGAGTGAAGAAGGTTTTCGGATCGTAAAACTCTGTTGTTAGAGAAGAACAAGGATGAGAGTAACTGTTCATCCCTTGACGGTATCTAACCAGAAAGCCACGGCTAACTACGTGCCAGCAGCCGCGGTAATACGTAGGTGGCAAGCGTTGTCCGGATTTATTGGGCGTAAAGCGAGCGCAGGCGGTTTCTTAAGTCTGATGTGAAAGCCCCCGGCTCAACCGGGGAGGGTCATTGGAAACTGGGAGACTTGAGTGCAGAAGAGGAGAGTGGAATTCCATGTGTAGCGGTGAAATGCGTAGATATATGGAGGAACACCAGTGGCGAAGGCGACTCTCTGGTCTGTAACTGACGCTGAGGCTCGAAAGCGTGGGGAGCAAACAGG | pt___00054 | Enterococcus thailandicus | 417368 |
| TGGGGAATATTGCGCAATGGGGGAAACCCTGACGCAGCAACGCCGCGTGAGCGATGAAGGTTTTCGGATCGTAAAGCTCTGTCTTTGGGGAAGATAATGACGGTACCCAAGGAGGAAGCTCCGGCTAACTACGTGCCAGCAGCCGCGGTAATACGTAGGGAGCAAGCGTTGTCCGGATTCACTGGGCGTAAAGAGCACGTAGGCGGTTAATTAAGTCAGGTGTGAAAGTTTTCGGCTCAACCGGAAAAGTGCACTTGAAACTGAATAACTTGAGTATCGGAGAGGTAAGCGGAATTCCTAGTGTAGCGGTGAAATGCGTAGAGATTAGGAAGAACACCGGTGGCGAAGGCGGCTTACTGGACGATAACTGACGCTGAGGTGCGAAAGCGTGGGGAGCGAACAGG | pt___00055 | Anaerofustis stercorihominis | 214853 |
| TAGGGAATCTTCGGCAATGGGGGCAACCCTGACCGAGCAACGCCGCGTGAGTGAAGAAGGTTTTCGGATCGTAAAGCTCTGTTGTAAGAGAAGAACGAGTGTGAGAGTGGAAAGTTCACGCTGTGACGGTATCTTACCAGAAAGGGACGGCTAACTACGTGCCAGCAGCCGCGGTAATACGTAGGTCCCGAGCGTTATCCGGATTTATTGGGCGTAAAGCGAGCGCAGGCGGTTAGATAAGTCTGAAGTTAAAGGCTGTGGCTTAACCATAGTACGCTTTGGAAACTGTTTAACTTGAGTGCAAGAGGGGAGAGTGGAATTCCATGTGTAGCGGTGAAATGCGTAGATATATGGAGGAACACCGGTGGCGAAAGCGGCTCTCTGGCTTGTAACTGACGCTGAGGCTCGAAAGCGTGGGGAGCAAACAGG | pt___00055 | Streptococcus peroris | 68891 |
| TGGGGAATATTGCACAATGGGCGAAAGCCTGATGCAGCAACGCCGCGTGAGCGATGAAGGCCTTCGGGTCGTAAAGCTCTGTCCTCAAGGAAGATAATGACGGTACTTGAGGAGGAAGCCCCGGCTAACTACGTGCCAGCAGCCGCGGTAATACGTAGGGGGCTAGCGTTATCCGGAATTACTGGGCGTAAAGGGTGCGTAGGCGGTCTTTCAAGTCAGGAGTGAAAGGCTACGGCTCAACCGTAGTAAGCTCTTGAAACTGTAAGACTTGAGTGCAGGAGAGGAGAGTAGAATTCCTAGTGTAGCGGTGAAATGCGTAGATATTAGGAGGAATACCAGTTGCGAAGGCGGCTCTCTGGACTGTAACTGACGCTGAGGCACGAAAGCGTGGGGAGCAAACAGG | pt___00057 | Peptostreptococcaceae | 186804 |
| TGAGGAATATTGGTCAATGGCCGAGAGGCTGAACCAGCCAAGTCGCGTGAAGGATGAAGGATCTATGGTTTGTAAACTTCTTTTATAGGGGAATAAAGTGTGGGACGTGTTCCATTTTGTATGTACCCTATGAATAAGCATCGGCTAACTCCGTGCCAGCAGCCGCGGTAATACGGAGGATGCGAGCGTTATCCGGATTTATTGGGTTTAAAGGGTGCGTAGGTGGTAATTTAAGTCAGCGGTGAAAGTTTGTGGCTCAACCATAAAATTGCCGTTGAAACTGGGTTACTTGAGTGTGTTTGAGGTAGGCGGAATGCGTGGTGTAGCGGTGAAATGCATAGATATCACGCAGAACTCCAATTGCGAAGGCAGCTTACTAAACCATAACTGACACTGAAGCACGAAAGCGTGGGTATCAAACAGG | pt___00067 | Parabacteroides johnsonii | 387661 |
| TGAGGAATATTGGTCAATGGCCGAGAGGCTGAACCAGCCAAGTCGCGTGAAGGAAGAAGGATCTATGGTTTGTAAACTTCTTTTATAGGGGAATAAAGTGGAGGACGTGTCCTTTTTTGTATGTACCCTATGAATAAGCATCGGCTAACTCCGTGCCAGCAGCCGCGGTAATACGGAGGATGCGAGCGTTATCCGGATTTATTGGGTTTAAAGGGTGCGTAGGTGGTGATTTAAGTCAGCGGTGAAAGTTTGTGGCTCAACCATAAAATTGCCGTTGAAACTGGGTTACTTGAGTGTGTTTGAGGTAGGCGGAATGCGTGGTGTAGCGGTGAAATGCATAGATATCACGCAGAACTCCGATTGCGAAGGCAGCTTACTAAACCATAACTGACACTGAAGCACGAAAGCGTGGGGATCAAACAGG | pt___00067 | Parabacteroides merdae | 46503 |
| TGGGGAATATTGCACAATGGAGGAAACTCTGATGCAGCGACGCCGCGTGAAGGATGAAGTATTTCGGTATGTAAACTTCTATCAGCAGGGAAGAAAATGACGGTACCTGACTAAGAAGCCCCGGCTAACTACGTGCCAGCAGCCGCGGTAATACGTAGGGGGCAAGCGTTATCCGGATTTACTGGGTGTAAAGGGAGCGTAGACGGCACGGCAAGCCAGATGTGAAAGCCCGGGGCTCAACCCCGGGACTGCATTTGGAACTGCTGAGCTAGAGTGTCGGAGAGGCAAGTGGAATTCCTAGTGTAGCGGTGAAATGCGTAGATATTAGGAGGAACACCAGTGGCGAAGGCGGCTTGCTGGACGATGACTGACGTTGAGGCTCGAAAGCGTGGGGAGCAAACAGG | pt___00068 | Dorea longicatena | 88431 |
| TAGGGAATCTTCCGCAATGGACGCAAGTCTGACGGAGCAACGCCGCGTGAGTGAAGAAGGATTTCGGTTCGTAAAACTCTGTTGTTAGAGAAGAACAGCGCATAGAGTAACTGTTATGCGTGTGACGGTATCTAACCAGAAAGCCACGGCTAACTACGTGCCAGCAGCCGCGGTAATACGTAGGTGGCAAGCGTTGTCCGGATTTATTGGGCGTAAAGCGAGCGCAGGCGGTCAATTAAGTCTGATGTGAAAGCCCCCGGCTCAACCGGGGAGGGTCATTGGAAACTGGTTGACTTGAGTGCAGAAGAGGAGAGTGGAATTCCATGTGTAGCGGTGAAATGCGTAGATATATGGAGGAACACCAGTGGCGAAGGCGACTCTCTGGTCTGTAACTGACGCTGAGGCTCGAAAGCGTGGGTAGCAAACAGG | pt___00074 | Granulicatella elegans | 137732 |
| TAGGGAATCTTCCACAATGGACGAAAGTCTGATGGAGCAACGCCGCGTGAGTGAAGAAGGTTTTCGGATCGTAAAGCTCTGTTGTTGGTGAAGAAGGATAGAGGTAGTAACTGGCCTTTATTTGACGGTAATCAACCAGAAAGTCACGGCTAACTACGTGCCAGCAGCCGCGGTAATACGTAGGTGGCAAGCGTTGTCCGGATTTATTGGGCGTAAAGCGAGCGCAGGCGGAAGAATAAGTCTGATGTGAAAGCCCTCGGCTTAACCGAGGAACTGCATCGGAAACTGTTTTTCTTGAGTGCAGAAGAGGAGAGTGGAATTCCATGTGTAGCGGTGGAATGCGTAGATATATGGAAGAACACCAGTGGCGAAGGCGACTCTCTGGTCTGCAACTGACGCTGAGGCTCGAAAGCATGGGTAGCGAACAGG | pt___00074 | Lactobacillus helveticus | 1587 |

| | | | |
|---|---|---|---|
| TGAGGAATATTGGTCAATGGACGCAAGTCTGAACCAGCCATGCCGCGTGCAGGAAGACGGCTCTATGAGTTGTAAACTGCTTTTGTACGAGGGTAACAGCATCTACGCGTAGGTGCATGAAAGTATCGTACGAATAAGGATCGGCTAACTCCGTGCCAGCAGCCGCGGTAATACGGAGGATCCAAGCGTTATCCGGATTTATTGGGTTTAAAGGGTGCGTAGGCGGTTTTATAAGTTAGAGGTTAAATATCGGAGCTTAACTCCGTTATGCCTCTAATACTGTAGGACTAGAGAATAGTTGCGGTAGGCGGAATGTATGGTGTAGCGGTGAAATGCTTAGAGATCATACAGAACACCGATTGCGAAGGCAGCTTACCAAACTATGACTGACGTTGAGGCACGAAAGCGTGGGGAGCAAACAGG | pt___00097 | Alistipes | 239759 |
| GGGGAATATTGGGCAATGGGCGCAAGCCTGACCCAGCAACGCCGCGTGAAGGAAGAAGGCTTTCGGGTTGTAAACTTCTTTTGTCAGGGACGAAACAAATGACGGTACCTGACGAATAAGCCACGGCTAACTACGTGCCAGCAGCCGCGATAATACGTAGGGGGCTAGCGTTATCCGGAATTACTGGGCGTAAAGGGTGCGTAGGCGGTCTTTCAAGCCAGAAGTGAAAGGCTACGGCTCAACCGTAGTAAGCTTTTGGAACTGTAGGACTTGAGTGCAGGAGAGGAGAGTGGAATTCCTAGTGTAGCGGTGAAATGCGTAGATATTAGGAGGAACACCAGTAGCGAAGGCGGCTCTCTGGACTGTAACTGACGCTGAGGCACGAAAGCGTGGGGAGCAAACAGG | pt___00097 | Paeniclostridium ghonii | 29358 |
| TGGGGAATATTGCACAATGGGCGAAAGCCTGATGCAGCGACGCCGCGTGAGTGAAGAAGTATTTCGGTATGTAAAGCTCTATCAGCAGGGAAGATAATGACAGTACCTGACTAAGAAGCCCCGGCTAACTACGTGCCAGCAGCCGCGGTAATACGTAGGGGGCAAGCGTTATCCGGATTTACTGGGTGTAAAGGGAGCGTAGGTGGCTAGGTAAGTCAGGTGTGAAAGCCCGGGGCTCAACCCCGGGATTGCACTTGAAACTACTTAGCTAGAGTGCAGGAGAGGTAAGTGGAATTCCTAGTGTAGCGGTGAAATGCGTAGATATTAGGAGGAACACCAGTGGCGAAGGCGGCTTACTGGACTGTAACTGACACTGAGGCTCGAAAGCGTGGGGAGCAAACAGG | pt___00111 | Mobilitalea sibirica or [Clostridium] fimetarium | 1462919 or 99656 |
| TGGGGAATATTGCACAATGGGGGAAACCCTGATGCAGCGACGCCGCGTGAGCGAAGAAGTATTTCGGTATGTAAAGCTCTATCAGCAGGGAAGAAGAAATGACGGTACCTGACTAAGAAGCACCGGCTAAATACGTGCCAGCAGCCGCGGTAATACGTATGGTGCAAGCGTTATCCGGATTTACTGGGTGTAAAGGGAGCGCAGGCGGAAGGCTAAGTCTGATGTGAAAGCCCGGGGCTCAACCCCGGTACTGCATTGGAAACTGGTCATCTAGAGTGTCGGAGGGGTAAGTGGAATTCCTAGTGTAGCGGTGAAATGCGTAGATATTAGGAGGAACACCAGTGGCGAAGGCGGCTTACTGGACGATAACTGACGCTGAGGCTCGAAAGCGTGGGGAGCAAACAGG | pt___00117 | Roseburia inulinivorans | 360807 |
| TGGGGAATATTGCACAATGGGCGCAAGCCTGATGCAGCCATGCCGCGTGTATGAAGAAGGCCTTCGGGTTGTAAAGTACTTTCAGTCGGGAGGAAGGTGTCAAGGTTAATAACCTTGGCAATTGACGTTACCGACAGAAGAAGCACCGGCTAACTCCGTGCCAGCAGCCGCGGTAATACGGAGGGTGCAAGCGTTAATCGGAATTACTGGGCGTAAAGCGCACGCAGGCGGTTGATTGAGTCAGATGTGAAATCCCCGGGCTTAACCCGGGAATTGCATCTGATACTGGTCAGCTAGAGTCTTGTAGAGGGGGGTAGAATTCCATGTGTAGCGGTGAAATGCGTAGAGATGTGGAGGAATACCGGTGGCGAAGGCGGCCCCCTGGACAAAGACTGACGCTCAGGTGCGAAAGCGTGGGGAGCAAACAGG | pt___00131 | Morganella morganii | 582 |
| TAGGGAATCTTTCACAATGGGCGAAAGCCTGATGGAGCAATGCCGCGTGCAGGACGAAGGCCTTCGGGTTGTAAACTGCTTTTAAAGCCGAGAAATATGATGGTAAGCTTTGAATAAGGATCGGCTAACTACGTGCCAGCAGCCGCGGTCATACGTAGGATCCGAGCATTATCCGGAGTGACTGGGTGTAAAGAGTTGCGTAGGTGGCAAGGTAAGTAGATAGTGAAATCTGGTGGCTCAACCATTCAGACTATTATCTAAACTATCTAGCTCGAGACTGTTATGGGTAACTGGAATTTCTAGTGTAGGAGTGAAATCCGTAGATATTAGAAGGAACACCAATAGCGTAGGCAGGTTACTGGAACAGTTCTGACACTAAGGCACGAAAGCGTAGGGAGCAAACGGG | pt___00131 | Candidatus Saccharibacteria | 95818 |
| TGAGGAATATTGGTCAATGGACGGAAGTCTGAACCAGCCAAGTAGCGTGCAGGATGACGGCCCTATGGGTTGTAAACTGCTTTTGTATGGGGATAAAGTTAGGGACGTGTCCCTATTTGCAGGTACCATACGAATAAGGACCGGCTAATTCCGTGCCAGCAGCCGCGGTAATACGGAAGGTCCAGGCGTTATCCGGATTTATTGGGTTTAAAGGGAGCGTAGGCTGGATATTAAGTGTGTTGTGAAATGTAGACGCTCAACGTCTGACTTGCAGCGCATACTGGTTTCCTTGAGTACGCACAACGTTGGCGGAATTCGTCGTGTAGCGGTGAAATGCTTAGATATGACGAAGAACTCCGATTGCGAAGGCAGCTGACGGGAGCGCAACTGACGCTTAAGCTCGAAGGTGCGGGTATCAAACAGG | pt___00142 | Prevotella | 838 |
| TGAGGAATATTGGTCAATGGACGCAAGTCTGAACCAGCCATGCCGCGTGCAGGATGAAGGTGCTATGCATTGTAAACTGCTTTTGTACGAGGGTAAATGCAGGTACGTGTACCTGTTTGAAAGTATCGTACGAATAAGGGTCGGCTAACTCCGTGCCAGCAGCCGCGGTAATACGGAGGACCCGAGCGTTATCCGGATTTATTGGGTTTAAAGGGTGCGTAGGCGGATTAGTAAGTTAGAGGTGAAAGCTCGATGCTCAACATCGAAATTGCCTCTGATACTGTTAGTCTAGAGTATAGTTGCGGAAGGCGGAATGTGTGGTGTAGCGGTGAAATGCTTAGATATCACACAGAACACCGATTGCGAAGGCAGCTTTCCAAGCTATTACTGACGCTGATGCACGAAAGCGTGGGGAGCGAACAGG | pt___00150 | Alistipes indistinctus | 626932 |
| TAGGGAATCTTCCACAATGGACGCAAGTCTGATGGAGCAACGCCGCGTGAGTGAAGAAGGTCTTCGGATCGTAAAGCTCTGTTGTTGGTGAAGAAGGATAGAGGCAGTAACTGGTCTTTATTTGACGGTAATCAACCAGAAAGTCACGGCTAACTACGTGCCAGCAGCCGCGGTAATACGTAGGTGGCAAGCGTTGTCCGGATTTATTGGGCGTAAAGCGAGCGCAGGCGGAATGATAAGTCTGATGTGAAAGCCCACGGCTCAACCGTGGAACTGCATCGGAAACTGTCATTCTTGAGTGCAGAAGAGGAGAGTGGAACTCCATGTGTAGCGGTGGAATGCGTAGATATATGGAAGAACACCAGTGGCGAAGGCGGCTCTCTGGTCTGCAACTGACGCTGAGGCTCGAAAGCATGGGTAGCGAACAGG | pt___00157 | Lactobacillus delbrueckii | 1584 |
| TGGGGAATATTGCACAATGGAGGAAACTCTGATGCAGCGACGCCGCGTGAGGGAAGAAGGTCTTCGGATTGTAAACCTCTGTCTTCAGGGACGATAATGACGGTACCTGAGGAGGAAGCCACGGCTAACTACGTGCCAGCAGCCGCGGTAAAACGTAGGTGGCAAGCGTTGTCCGGAATTACTGGGTGTAAAGGGAGTGCAGGCGGGACGGCAAGTTGGAAGTGAAACCCATGGGCTTAACCCATGAACTGCTTTCAAAACTGTCGTTCTTGAGTGGTGCAGAGGTAGGCGGAATTCCCGGTGTAGCGGTGGAATGCGTAGATATCGGGAGGAACACCAGTGGCGAAGGCGGCCTACTGGGCACTAACTGACGCTGAGGCTCGAAAGCATGGGTAGCAAACAGG | pt___00228 | Eubacteriales | 186802 |

| | | | |
|---|---|---|---|
| TGGGGAATATTGCACAATGGGGGAAACCCTGATGCAGCAACGCCGCGTGAGTGAAGAAGTATTTCGGTATGTAAAGCTCTA TCAGCAGGGAAGATAATGACGGTACCTGACTAAGAAGCTCCGGCTAAATACGTGCCAGCAGCCGCGGTAATACGTATGGAG CAAGCGTTATCCGAATTTACTGGGTGTAAAGGGTGCGTAGGTGGCAGTGCAAGTCAGATGTGAAAGGCCGGGGCTCAACCC CGGAGCTGCATTTGAAACTGCATAGCTAGAGTACAGGAGAGGCAGGCGGAATTCCTAGTGTAGCGGTGAAATGCGTAGATA TTAGGAGGAACACCAGTGGCGAAGGCGGCCTGCTGGACTGTTACTGACACTGAGGCACGAAAGCGTGGGGAGCAAACAGG | pt___00421 | Lachnospiraceae | 186803 |
| TGGGGAATATTGGACAATGGGGGGAACCCTGATCCAGCCATGCCGCGTGTGTGAAGAAGGCCTTTTGGTTGTAAAGCACTT TAAGCGAGGAGGAGGCTTACCTGGTTAATACCTGGGATAAGTGGACGTTACTCGCAGAATAAGCACCGGCTAACTCTGTGC CAGCAGCCGCGGTAATACAGAGGGTGCAAGCGTTAATCGGATTTACTGGGCGTAAAGCGCGCGTAGGTGGCTAATTAAGTC AAATGTGAAATCCCCGAGCTTAACTTGGGAATTGCATTCGATACTGGTTAGCTAGAGTATGGGAGAGGATGGTAGAATTCC AGGTGTAGCGGTGAAATGCGTAGAGATCTGGAGGAATACCGATGGCGAAGGCAGCCATCTGGCCTAATACTGACACTGAGG TGCGAAAGCATGGGGAGCAAACAGG | pt___00542 | Acinetobacter variabilis or Acinetobacter boissieri | 70346 or 1219383 |

Table ST6: Sequences of V3-V4 segment of rRNA gene that defined particular phylotypes included in ratios to describe compositional attributes with classified taxonomy and NCBI taxonomy identification. Phylogenetic classifications that could have been placed near two taxonomies were described with ’or’ designation.

### 4 Supplemental Material References

[7] James R. Cole, Qiong Wang, Jordan A. Fish, Benli Chai, Donna M. McGarrell, Yanni Sun, C. Titus Brown, Andrea Porras-Alfaro, Cheryl R. Kuske, and James M. Tiedje. Ribosomal database project: data and tools for high throughput rRNA analysis. *Nucleic Acids Research*, 42(D1):D633–D642, nov 2013.

[8] Alexandros Stamatakis. RAxML version 8: a tool for phylogenetic analysis and post-analysis of large phylogenies. *Bioinformatics*, 30(9):1312–1313, jan 2014.

[9] Samuel S Minot, Bailey Garb, Alennie Roldan, Alice Tang, Tomiko Oskotsky, Christopher Rosenthal, Noah G Hoffman, Marina Sirota, and Jonathan L Golob. Robust harmonization of microbiome studies by phylogenetic scaffolding with maliampi. *bioRxiv*, 2022.

[10] Jonathan L. Golob, Elisa Margolis, Noah G. Hoffman, and David N. Fredricks. Evaluating the accuracy of amplicon-based microbiome comp10.1093/nar/gkt1244utational pipelines on simulated human gut microbial communities. *BMC Bioinformatics*, 18(1), may 2017.

[11] Bastiaan W. Haak, Eric R. Littmann, Jean-Luc Chaubard, Amanda J. Pickard, Emily Fontana, Fatima Adhi, Yangtsho Gyaltshen, Lilan Ling, Sejal M. Morjaria, Jonathan U. Peled, Marcel R. van den Brink, Alexander I. Geyer, Justin R. Cross, Eric G. Pamer, and Ying Taur. Impact of gut colonization with butyrate producing microbiota on respiratory viral infection following allo-HCT. *Blood*, pages blood–2018–01–828996, apr 2018.

[12] Ying Taur, Joao B. Xavier, Lauren Lipuma, Carles Ubeda, Jenna Goldberg, Asia Gobourne, Yeon Joo Lee, Krista A. Dubin, Nicholas D. Socci, Agnes Viale, Miguel-Angel Perales, Robert R. Jenq, Marcel R. M. van den Brink, and Eric G. Pamer. Intestinal domination and the risk of bacteremia in patients undergoing allogeneic hematopoietic stem cell transplantation. *Clinical Infectious Diseases*, 55(7):905–914, jun 2012.

[13] Jonathan U. Peled, Antonio L.C. Gomes, Sean M. Devlin, Eric R. Littmann, Ying Taur, Anthony D. Sung, Daniela Weber, Daigo Hashimoto, Ann E. Slingerland, John B. Slingerland, Molly Maloy, Annelie G. Clurman, Christoph K. Stein-Thoeringer, Kate A. Markey, Melissa D. Docampo, Marina Burgos da Silva, Niloufer Khan, André Gessner, Julia A. Messina, Kristi Romero, Meagan V. Lew, Amy Bush, Lauren Bohannon, Daniel G. Brereton, Emily Fontana, Luigi A. Amoretti, Roberta J. Wright, Gabriel K. Armijo, Yusuke Shono, Míriam Sanchez-Escamilla, Nerea Castillo Flores, Ana Alarcon Tomas, Richard J. Lin, Lucrecia Yáñez San Segundo, Gunjan L. Shah, Christina Cho, Michael Scordo, Ioannis Politikos, Kasumi Hayasaka, Yuta Hasegawa, Boglarka Gyurkocza, Doris M. Ponce, Juliet N. Barker, Miguel-Angel Perales, Sergio A. Giralt, Robert R. Jenq, Takanori Teshima, Nelson J. Chao, Ernst Holler, Joao B. Xavier, Eric G. Pamer, and Marcel R.M. van den Brink. Microbiota as predictor of mortality in allogeneic hematopoietic-cell transplantation. *New England Journal of Medicine*, 382(9):822–834, feb 2020.
